# Tolerance for Adverse Events from Operative and Nonoperative Treatment for Mild Cervical Spondylotic Myelopathy

**DOI:** 10.64898/2026.08.21.26361046

**Authors:** Faraz Arkam, Eliana Goldstein, Xianqi Zeng, Salim Yakdan, Andrew K. Chan, Abby L. Cheng, Dean Chou, Matthew W. Colman, Zoher Ghogawala, Jakub Godzik, Michael P. Kelly, Thomas E. Mroz, Brian J. Neuman, Lindsay Orosz, Paul Park, Alpesh A. Patel, Eric A. Potts, John Rhee, Rick C. Sasso, Kenneth B. Schechtman, Michael P. Steinmetz, Grace X. Xiong, Linying Zhang, Wilson Z. Ray, Jacob K. Greenberg, Mary C. Politi

**Affiliations:** Department of Neurological Surgery, Washington University School of Medicine, St. Louis, Missouri, USA; Bursky School of Public Health, Washington University in St. Louis, St. Louis, Missouri, USA; Department of Neurological Surgery, Columbia University Vagelos College of Physicians and Surgeons, New York, USA; Department of Orthopaedic Surgery, Washington University School of Medicine, St. Louis, Missouri, USA; Department of Neurologic and Orthopaedic Surgery, Northwestern University Feinberg School of Medicine, Chicago, Illinois, USA; Department of Neurosurgery, Lahey Hospital and Medical Center, Burlington, Massachusetts, USA; Department of Neurosurgery, University of Alabama at Birmingham, Birmingham, Alabama, USA; Department of Orthopaedic Surgery, Stanford University School of Medicine, Stanford, California, USA; Department of Neurosurgery, Cleveland Clinic Lerner College of Medicine at Case Western Reserve University, Cleveland, Ohio, USA; National Spine Health Foundation, Reston, Virginia, USA; Semmes Murphey Clinic, Memphis, Tennessee, USA; Goodman Campbell Brain and Spine, Carmel, Indiana, USA; Department of Orthopaedic Surgery, Emory University School of Medicine, Atlanta, Georgia, USA; Department of Orthopaedic Surgery, Indiana University School of Medicine, Carmel, Indiana, USA; Division of Biostatistics, Washington University School of Medicine, St. Louis, Missouri, USA

## Abstract

**Background:** Guidelines recommend surgery for moderate and severe cervical spondylotic myelopathy (CSM) but support either surgery or nonoperative care for mild disease. Patients’ concerns about adverse events associated with each pathway can influence care choices, but these concerns are not well-studied.

**Methods:** We conducted a three-arm randomized vignette experiment among United States adults aged 40 years and older recruited through an online research panel. All participants read an identical description of mild CSM and were randomized to one of three scenarios: surgery that improved symptoms, surgery that halted progression without improvement, or nonoperative management with symptom progression. Participants in the surgical scenarios rated 12 possible complications and those in the nonoperative scenario rated 8 progression outcomes. For each item, participants rated how strongly each complication or progression outcome would influence their decision (0-10) and whether they would still choose the same care plan given the outcomes. Items for which participants would no longer choose the same treatment were termed “dominant decision factors.”

**Results:** Of 263 participants analyzed, 58% were female and 62% were aged 70 years or older. Median relative importance ratings ranged from 3 to 10 across the twelve surgical complications and from 5 to 8 across the eight progression outcomes. Complications were rated as more likely to influence the treatment decision when surgery was described as halting progression rather than improving symptoms (mean difference 0.45 to 1.44 points). The most common dominant decision factor was new neck pain persisting at one year (78% in Scenario 1, 88% in Scenario 2); complications resolving within three months were the least common (14% to 29%). When the same complication persisted at one year rather than resolving within three months, rates rose from 25% to 68% for dysphagia, 15% to 49% for hoarseness, and 20% to 72% for shoulder weakness. In the nonoperative scenario, the most common dominant decision factors were worsening bladder control (57%) and neck pain interfering with sleep (53%). Many participants were undecided (4% to 36% in the surgical scenarios, 27% to 43% in the nonoperative scenario).

**Conclusions:** Treatment decisions for mild CSM may be driven by the expected long-term nature of adverse events and their anticipated impact on daily quality of life, rather than by conventional neurological metrics or the described benefits of surgery.

## Introduction

Cervical spondylotic myelopathy (CSM) is the most common cause of spinal cord dysfunction in adults worldwide [1,2], with about 2% of adults affected in their lifetime [3,4]. Age-related degeneration narrows the cervical canal and compresses the cord, producing impaired dexterity, gait imbalance and sensory disturbance [5]. CSM can be classified as mild, moderate, or severe, graded using the modified Japanese Orthopaedic Association (mJOA) scale [6].

Treatment decisions and outcomes for CSM vary by disease severity. Among patients with moderate to severe disease, surgery often improves mJOA and quality of life scores at 12 months. Patients with mild CSM can achieve clinically important positive changes in quality of life (EQ-5D) and physical function (SF-36 PCS) after surgery [7], but they may show more modest neurological improvement than patients with moderate or severe disease [7]. Among patients with mild CSM initially treated without surgery, a sizable minority (around 12% in 12 months) choose to have surgery over time as symptoms progress or fail to improve. [8]

Surgical risk adds additional complexity to treatment decisions in patients with mild CSM. Perioperative complications occur in 11% to 38% of patients undergoing surgery for CSM, most commonly dysphagia, dural tear and superficial wound infection [6]. Most complications are transient, but some are not. Among patients who deteriorate neurologically after surgery for CSM, 48% still have a deficit of at least one mJOA point one year later [9]. Guidelines therefore recommend surgery for moderate and severe CSM but suggest patient-engaged or shared decision making between surgery or structured nonoperative care for mild disease [10].

There is sparse research regarding how patients weigh potential adverse events from CSM treatment, including how this affects their interest in surgery or nonoperative management. Both operative and nonoperative treatment carry risks, and patient tolerance for adverse events may vary depending on the degree of expected benefit from surgery.

We therefore conducted a three-arm randomized vignette experiment among United States adults aged 40 years and over, asking respondents to imagine the vignettes and associated surgical complications. The goal was to examine which surgical complications would influence treatment decisions and satisfaction.

## Methods

### Design and oversight

We conducted a three-arm, parallel-group, randomized vignette experiment administered as an anonymous online survey. The vignettes and survey instruments were developed by the study team, which included a combination of CSM clinical experts, comparative effectiveness researchers, and patient engagement and decision science experts. Scenario wording was informed by the clinical literature on complications following cervical decompression and by clinical experience in the management of CSM. All outcomes were described in lay terms, and the instrument was reviewed by clinical CSM and survey experts within the study team to assess clarity, relevance, and content.

The protocol was approved as exempt research by the Washington University in St. Louis Human Research Protection Office (IRB #202511100). All respondents were shown a consent information sheet electronically at the start of the survey. If they agreed to continue, they were shown the survey items. No identifiable health information was collected. Methods and results are reported in accordance with the Checklist for Reporting Results of Internet E-Surveys (CHERRIES) [11]; the completed checklist is Supplementary File 1.

### Participants and recruitment

Respondents were recruited through CloudResearch Prime Panels over a 1-week period between 14 and 20 May 2026. Prime Panels is an online research platform that maintains a standing pool of volunteers who agree to complete surveys in exchange for compensation [12]. The survey was administered using Qualtrics (Qualtrics, Provo, UT) and was distributed through the panel.

Eligibility was restricted to adult United States residents aged 40 years or older. Recruitment closed once 90 completed responses per arm had been obtained, an *a priori* determined number based on broad sample size estimates and feasibility for this exploratory study. The final number of collected responses differed slightly because some responses were submitted simultaneously and were retained.

### Randomization and Scenarios

After consent, participants were randomized 1:1:1 to one of three scenarios. All respondents first read an identical stem asking them to imagine living with a set of symptoms consistent with mild CSM (Supplementary File 2). The description stated that some people choose surgery while others monitor their symptoms, and that there is no single correct choice.

Respondents then read one of three scenarios. In Scenario 1, they choose to undergo surgery, symptoms improve at one year, and a complication has occurred. In Scenario 2, they choose to undergo surgery, symptoms are stable rather than better, and the same complication has occurred. In Scenario 3, they decline surgery and symptoms worsen. Scenarios 1 and 2 were word-for-word identical apart from the clause describing the benefit obtained.

### Items and outcomes

Respondents in Scenarios 1 and 2 rated the same 12 complications. Those in Scenario 3 rated 8 progression outcomes. Four complications were described as resolving within three months, and eight as persistent at one year, serious, or requiring reoperation. The full instrument is Supplementary File 2.

Each item was presented separately and was followed by two questions. The first asked how strongly that outcome would influence their decision about surgery, rated from 0 (Would **not** influence my decision at all) to 10 (Would influence my decision strongly); we refer to this as the “relative importance” of the item. The second asked whether the respondent would still choose surgery (Scenarios 1-2) or nonoperative treatment (Scenario 3) answered as Yes, No, or Unsure.

A response of “No” to the second question for each item indicated that the respondent would change their treatment choice based on the described outcome; we call these dominant decision factors, or “dominant factors” for brevity. For each item, the dominant decision factor rate was the number of "No" responses divided by the total of "No", "Unsure" and "Yes" responses. “Unsure” responses were grouped with “Yes”, so the dominant factor rate is a conservative estimate. The question is directional: in the surgical scenarios (1 & 2) “No” means the respondent would decline surgery, whereas in the non-operative scenario (3) it means they would elect for surgery. Two respondent-level outcomes were derived: the number of complications through which a respondent would still accept surgery, and the percentage of items falling into each of the three response categories.

### Data quality and sample size

A single attention-check item was embedded in each scenario, asking respondents to identify which of four items is typically cold (fire, ice, boiling water, sunlight). Thirteen respondents (4.7%) did not select the correct answer and were excluded before analysis. Speeding was defined within arm as a completion time below 50% of that arm’s median. [13] Eighteen respondents (6.8%) met this definition and were retained. Speeding was not prespecified as an exclusion criterion, and the responses were otherwise complete. We performed complete case analysis, excluding a small amount (<1%) of missing data.

### Statistical analysis

Ordinal variables are summarized as median (interquartile range) and categorical variables as counts with percentages and Wilson score 95% confidence intervals. [14] Characteristics were compared across the three scenarios using chi-square or Fisher exact tests to confirm that randomization produced comparable groups.

Relative importance of items was compared between Scenario 1 and Scenario 2 for each complication using the Mann-Whitney U test. Dominant factor rates were compared by Fisher exact test, with Newcombe hybrid-score confidence intervals for between-arm differences. [15] Twelve comparisons were made for each outcome, and p values are not adjusted for multiplicity.

Because twelve parallel comparisons were made regarding surgical complications, a two-sided exact sign test was utilized to determine whether the difference between the scenarios acted in a consistent direction across items, rather than varying from complication to complication. For each respondent we calculated the percentage of their items on which they maintained their treatment described in the scenario called “decision stability”. This was compared between the pooled surgical scenarios and the non-operative scenario utilizing a Mann-Whitney U test, with rank-biserial correlation calculated for effect size.

Associations between participant characteristics and the two respondent-level outcomes were exploratory. All tests were two-sided with α = 0.05. Analyses were performed using R version 4.4.2.

## Results

### Respondents

A total of 276 respondents completed the survey. After exclusion of those who failed the embedded attention check and/or met exclusion criteria, 263 (95.2%) were analyzed. Of these, 89 were allocated to Scenario 1, 90 to Scenario 2, and 84 to Scenario 3. Overall, 153 (58.2%) were female, 163 (62.0%) were aged 70 years or older, 140 (53.2%) had Medicare insurance, and 27 (10.3%) reported prior spine surgery.

### Relative Importance of Adverse Events

Median relative importance ratings varied widely between complications. The lowest-rated item in each was a minor wound infection resolving within three months, with a median of 3 (IQR 1–6) in Scenario 1 and 4 (1–7) in Scenario 2. The highest-rated items were new neck pain persisting at one year, with a median of 10 (IQR 8–10) in both scenarios, and shoulder weakness persisting at one year, with a median of 9 (6–10) in Scenario 1 and 10 (7–10) in Scenario 2 (Supplementary Figure 1; Supplementary Table S1, Panel A).

Participants rated adverse events as more influential when surgery was described as halting progression (Scenario 2) rather than improving symptoms (Scenario 1). Mean ratings were higher in Scenario 2 for all twelve complications (exact sign test p < 0.001), but the differences were small, ranging from 0.45 to 1.44 points on the 0–10 scale. Seven of the twelve comparisons had p < 0.05 before adjustment for multiple comparisons. The largest difference was for transient shoulder weakness, whose median rose from 5 (IQR 3–7) to 7 (5–9) (p < 0.001).

In the nonoperative scenario, median relative importance ratings for the eight progression outcomes ranged from 5 (IQR 4-8) for increased difficulty handling small objects to 8 (5-9) for worsening bladder control (Supplementary Figure 2; Supplementary Table S1, Panel B).

### Dominant Decision Factors: Factors Influencing Surgery vs. Nonoperative Management

Complications described as persisting at one year, serious, or requiring reoperation were more often dominant decision factors than those resolving within three months. In Scenario 1, the most common dominant decision factor was new neck pain persisting at one year, rated a dominant factor by 77.5% of participants (95% CI 67.8–85.0), and the least common was hoarseness resolving within three months, at 13.5% (7.9–22.1). In Scenario 2, the most common was again new neck pain persisting at one year, at 87.8% (79.4–93.0), and the least common was a minor wound infection resolving within three months, at 15.7% (9.6–24.7).

In both scenarios, new neck pain persisting at one year was a dominant factor more often than hardware failure requiring reoperation (Scenario 1, 57.3%; Scenario 2, 65.6%), deep wound infection requiring reoperation (45.3%; 58.4%) and serious lung infection requiring reintubation (52.8%; 62.2%), with non-overlapping 95% confidence intervals (Table 2, Panel A). Dominant decision factor rates did not differ significantly between Scenario 1 and Scenario 2 for any complication, with differences ranging from −8.1 to +13.1 percentage points (Table 1, Panel A).

**Table 1.**
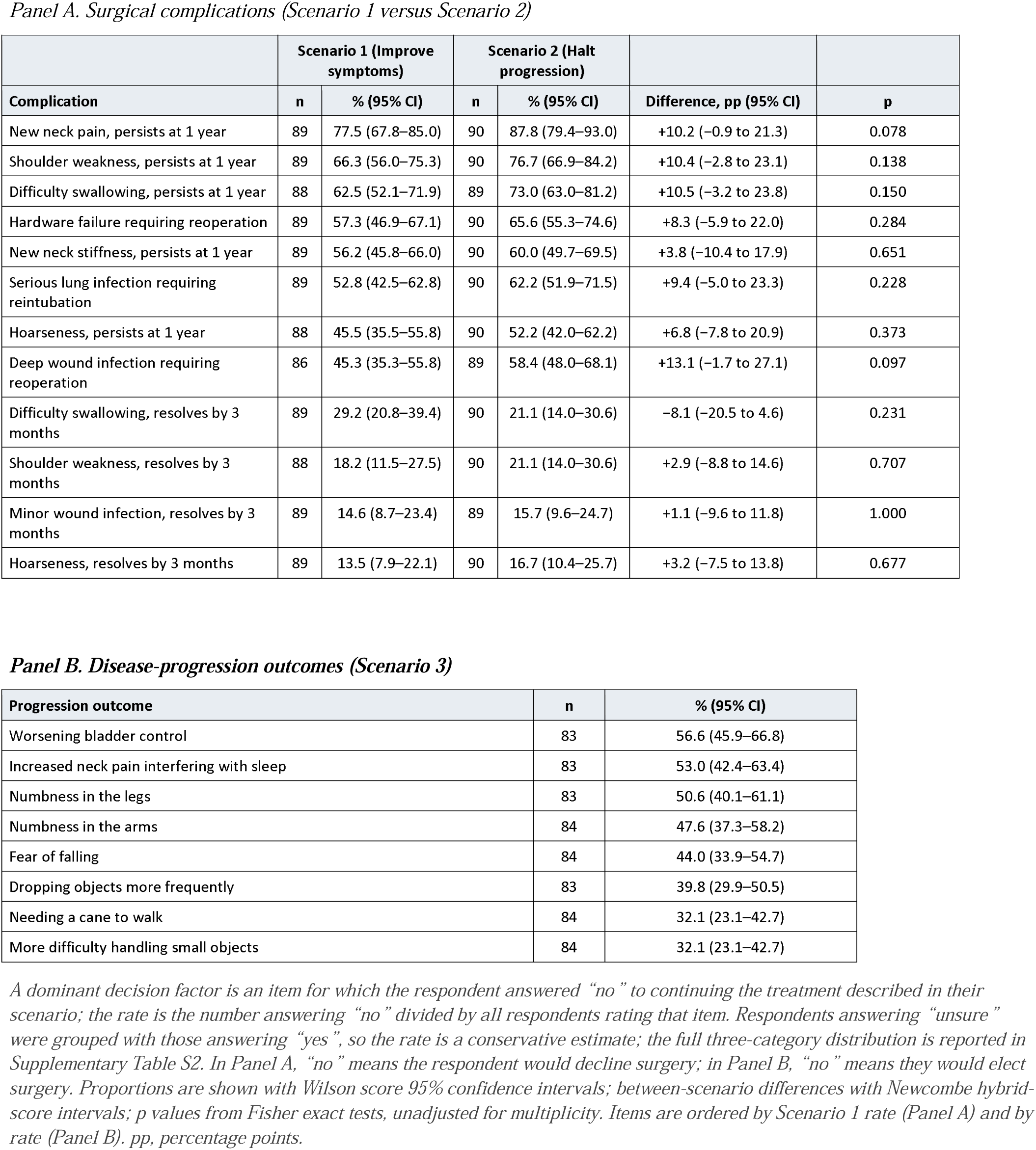
Dominant decision factor rates by adverse event and randomized scenario.

| *Panel A. Surgical complications (Scenario 1 versus Scenario 2)* |  |  |  |  |  |  |
|  | **Scenario 1 (Improve symptoms)** |  | **Scenario 2 (Halt progression)** |  |  |  |
| **Complication** | **n** | **% (95% CI)** | **n** | **% (95% CI)** | **Difference, pp (95% CI)** | **p** |
|---|---|---|---|---|---|---|
| New neck pain, persists at 1 year | 89 | 77.5 (67.8–85.0) | 90 | 87.8 (79.4–93.0) | +10.2 (−0.9 to 21.3) | 0.078 |
| Shoulder weakness, persists at 1 year | 89 | 66.3 (56.0–75.3) | 90 | 76.7 (66.9–84.2) | +10.4 (−2.8 to 23.1) | 0.138 |
| Difficulty swallowing, persists at 1 year | 88 | 62.5 (52.1–71.9) | 89 | 73.0 (63.0–81.2) | +10.5 (−3.2 to 23.8) | 0.150 |
| Hardware failure requiring reoperation | 89 | 57.3 (46.9–67.1) | 90 | 65.6 (55.3–74.6) | +8.3 (−5.9 to 22.0) | 0.284 |
| New neck stiffness, persists at 1 year | 89 | 56.2 (45.8–66.0) | 90 | 60.0 (49.7–69.5) | +3.8 (−10.4 to 17.9) | 0.651 |
| Serious lung infection requiring reintubation | 89 | 52.8 (42.5–62.8) | 90 | 62.2 (51.9–71.5) | +9.4 (−5.0 to 23.3) | 0.228 |
| Hoarseness, persists at 1 year | 88 | 45.5 (35.5–55.8) | 90 | 52.2 (42.0–62.2) | +6.8 (−7.8 to 20.9) | 0.373 |
| Deep wound infection requiring reoperation | 86 | 45.3 (35.3–55.8) | 89 | 58.4 (48.0–68.1) | +13.1 (−1.7 to 27.1) | 0.097 |
| Difficulty swallowing, resolves by 3 months | 89 | 29.2 (20.8–39.4) | 90 | 21.1 (14.0–30.6) | −8.1 (−20.5 to 4.6) | 0.231 |
| Shoulder weakness, resolves by 3 months | 88 | 18.2 (11.5–27.5) | 90 | 21.1 (14.0–30.6) | +2.9 (−8.8 to 14.6) | 0.707 |
| Minor wound infection, resolves by 3 months | 89 | 14.6 (8.7–23.4) | 89 | 15.7 (9.6–24.7) | +1.1 (−9.6 to 11.8) | 1.000 |
| Hoarseness, resolves by 3 months | 89 | 13.5 (7.9–22.1) | 90 | 16.7 (10.4–25.7) | +3.2 (−7.5 to 13.8) | 0.677 |
| ***Panel B. Disease-progression outcomes (Scenario 3)*** |  |  |  |  |  |  |
| **Progression outcome** | **n** | **% (95% CI)** |  |  |  |  |
| Worsening bladder control | 83 | 56.6 (45.9–66.8) |  |  |  |  |
| Increased neck pain interfering with sleep | 83 | 53.0 (42.4–63.4) |  |  |  |  |
| Numbness in the legs | 83 | 50.6 (40.1–61.1) |  |  |  |  |
| Numbness in the arms | 84 | 47.6 (37.3–58.2) |  |  |  |  |
| Fear of falling | 84 | 44.0 (33.9–54.7) |  |  |  |  |
| Dropping objects more frequently | 83 | 39.8 (29.9–50.5) |  |  |  |  |
| Needing a cane to walk | 84 | 32.1 (23.1–42.7) |  |  |  |  |
| More difficulty handling small objects | 84 | 32.1 (23.1–42.7) |  |  |  |  |

*A dominant decision factor is an item for which the respondent answered “no” to continuing the treatment described in their scenario; the rate is the number answering “no” divided by all respondents rating that item. Respondents answering “unsure” were grouped with those answering “yes”, so the rate is a conservative estimate; the full three-category distribution is reported in Supplementary Table S2. In Panel A, “no” means the respondent would decline surgery; in Panel B, “no” means they would elect surgery. Proportions are shown with Wilson score 95% confidence intervals; between-scenario differences with Newcombe hybrid-score intervals; p values from Fisher exact tests, unadjusted for multiplicity. Items are ordered by Scenario 1 rate (Panel A) and by rate (Panel B). pp, percentage points.*

Undecided responses ranged from 4.4% to 36.0% across both surgical scenarios. The undecided proportion differed significantly between scenarios for one complication only: new neck pain persisting at one year, from 14.6% in Scenario 1 to 4.4% in Scenario 2 (p = 0.023) (Figure 1; Supplementary Table S2, Panel A).

**Figure 1.**
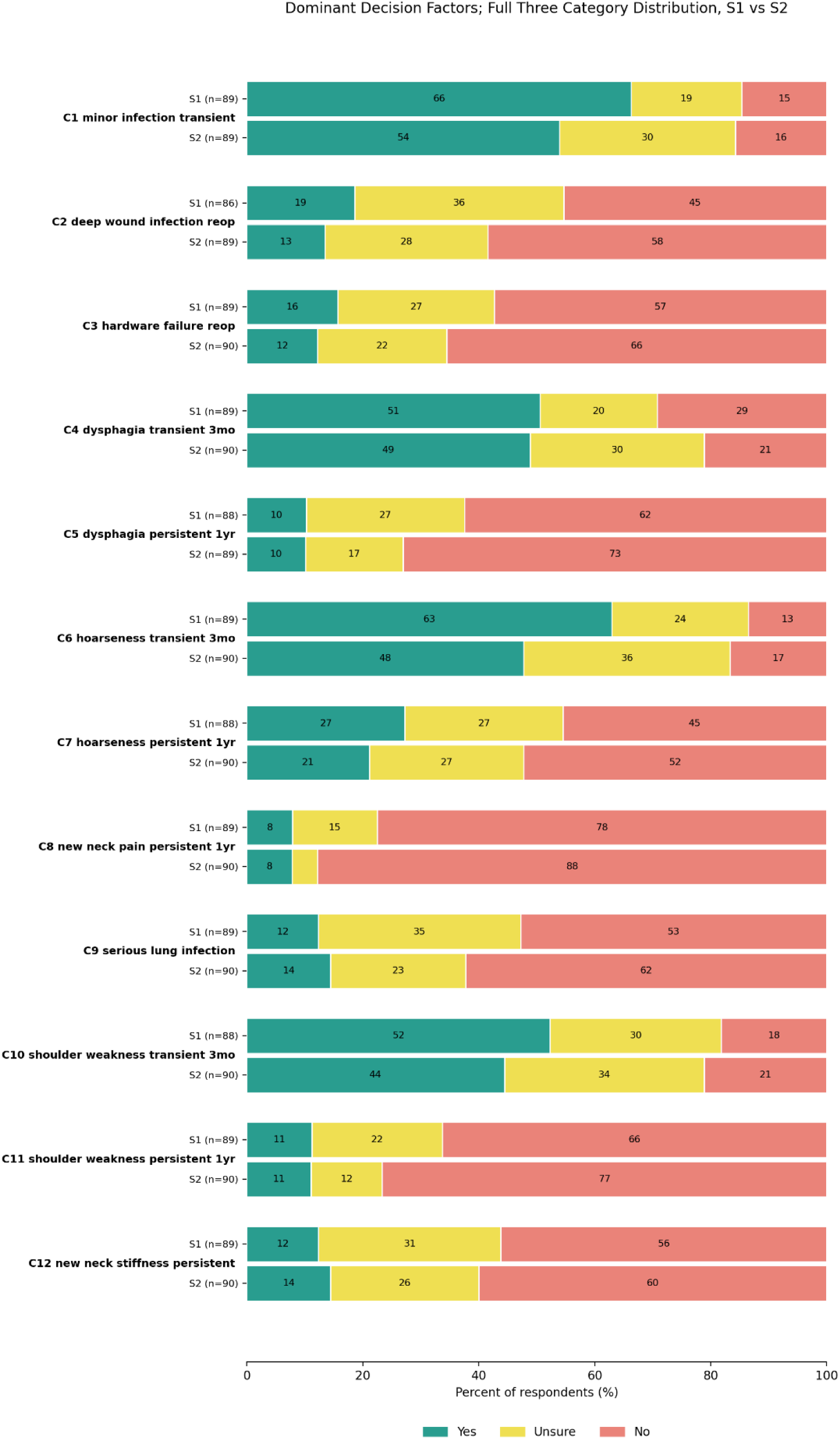
Treatment choice for each surgical complication, by randomized scenario. S1, surgery improves symptoms; S2, surgery halts progression. Bars show the percentage of respondents who would still choose surgery (Yes), were unsure (Unsure), or would decline surgery (No). The No segment is the dominant decision factor rate reported in Table 2, Panel A. C1–C12 correspond to the complications listed in Supplementary Table S1.

In Scenario 3, the most common dominant decision factors were worsening bladder control, identified by 56.6% of participants (95% CI 45.9–66.8), and increased neck pain interfering with sleep, at 53.0% (42.4–63.4); the least common were needing a cane to walk and increased difficulty handling small objects, each at 32.1% (23.1–42.7). Undecided responses ranged from 26.5% to 42.9%, the highest being for needing a cane to walk (Table 1, Panel B).

### Effect of the Described Duration of a Complication

Three complications were presented twice to respondents in the surgical scenarios, worded identically except for the stated duration. Because dominant decision factor rates did not differ significantly between scenarios for any complication, and because each respondent rated both versions, the two surgical scenarios were pooled for this within-person comparison. When the complication was described as persisting at one year rather than resolving within three months, dominant decision factor rates rose from 25.1% to 67.8% for dysphagia, from 15.1% to 48.9% for hoarseness, and from 19.7% to 71.5% for shoulder weakness. Further, for most complications resolving within three months, more participants were undecided than would change their treatment choice; the only exception was difficulty swallowing at 3 months in Scenario 1 (Supplementary Table S2, Panel A).

### Effect of participant characteristics

We explored the influence of participant demographic characteristics and history of surgery on outcomes. Age, sex, education and prior spine surgery were not associated with either the mean relative importance rating or the proportion of complications rated as dominant factors. Participants with Medicaid coverage (n=20) rated a smaller proportion of complications as dominant factors than participants with private insurance, a difference of 15.0 percentage points (p = 0.03).

## Discussion

In this randomized experiment, participants were asked to imagine that they had mild CSM and to consider whether they would choose or avoid surgery after a specific adverse outcome. Three main findings emerged. First, participants considering surgery were most influenced by the long-term persistence of adverse events. Second, participants considering non-operative treatment were most influenced by worsening bladder control and by neck pain. Third, participants who were told that surgery would halt progression rated every complication as more important to their decision than participants who were told that surgery would improve symptoms, though their stated treatment choices changed only marginally.

Duration was the strongest determinant of how important a complication was to the decision. Complications that resolved by 3 months were less influential than the same complications persisting at one year. This is clinically relevant, as permanent postoperative deficits are not uncommon in CSM. In a Canadian cohort of patients undergoing surgery for CSM, 50/428 (12%) deteriorated neurologically by three months, and 48% of those still had a deficit of at least one mJOA point at one year [9]. Higher baseline mJOA score was associated with deterioration, meaning patients with milder disease were more likely to experience this outcome [9]. Deterioration also occurs under nonoperative management: approximately 13-25% of patients with mild CSM experience significant decline by at least two mJOA points in the first year of nonoperative treatment [8]. Both treatment pathways therefore carry a risk of neurological decline, which patients must weigh alongside the adverse events specific to surgery.

New persistent neck pain at one year was the most common dominant decision factor in both surgical scenarios, above reoperation, deep wound infection, and a serious lung infection requiring reintubation. In the non-operative scenario, worsening bladder control and neck pain interfering with sleep were more often dominant decision factors than needing a cane to walk. Both findings suggest that symptoms expected to significantly impact quality of life were more influential than the neurological measures used to define disease severity. Therefore, while essential to measuring CSM outcomes, clinical measures of neurological abilities may not capture the real-world function or the outcomes most important to patients, such as sleep disturbance, which is common in this population but absent from the mJOA [17,21]. This conclusion is consistent with the use of quality-of-life measures, such as the SF-36, in recent CSM trials,[18] with documented disparities in patient-reported quality of life among patients with CSM [19] and results from recent patient surveys.[20]

Describing surgery as halting progression rather than improving symptoms modestly increased how much complications influenced the decision but did not significantly change treatment choice. A 0–10 rating may detect smaller shifts than a three-option choice question. One practical reading is that surgeons who describe a modest, expected benefit accurately are unlikely to deter patients who would otherwise choose surgery, though hypothetical survey choices may diverge from actual clinical decisions.

These findings have two practical implications. For patient communication, the expected duration of a complication should be stated explicitly: the same complication was roughly three times as likely to change a participant’s treatment choice when described as persisting at one year rather than resolving within three months. Decision-making and consent discussions that describe what a complication is, without saying how long it is expected to last, omit the attribute that mattered most in this study.

Providing such estimates will require long-term follow-up data on how often, and for how long, complications persist after surgery for mild CSM. For outcome measurement, the events that most often changed treatment choice were symptoms affecting day-to-day comfort, such as new persistent neck pain, worsening bladder control, and disturbed sleep, rather than the limb motor and sensory deficits used to grade disease severity. Trials in mild CSM therefore should pair mJOA with patient-reported measures that capture these symptoms, since a study powered on neurological change alone could miss the outcomes patients weigh most heavily.

### Limitations

This study has several limitations. Participants were members of the public asked to imagine a diagnosis, so their responses indicate hypothetical stated preferences rather than actual treatment decisions. Complications were presented one at a time, whereas a patient considering surgery faces the possibility of several occurring together. Relative importance ratings for the more severe complications were concentrated toward the upper end of the scale, limiting the ability to detect an effect related to the description of the surgical benefit. Responses of "Unsure" were grouped with "Yes," so the dominant factor rate is a conservative estimate and does not distinguish participants who were undecided from those who would proceed.

## Conclusion

Adults asked to imagine mild CSM based their stated treatment decisions largely on how long an adverse event was expected to last and how it would affect daily life. Describing surgery as halting progression rather than improving symptoms made participants rate every complication as more influential but changed stated decisions only slightly. Participants weighing nonoperative treatment were strongly influenced by the prospect of worsening symptoms, with worsening bladder control and neck pain being most influential. These findings may inform future studies evaluating adverse events and treatment decisions in mild CSM.

## Supporting information

Supplementary Material - Survey

## Data Availability

All data produced in the present study are available upon reasonable request to the authors

## Supplementary Tables

**Supplementary Table S1.**
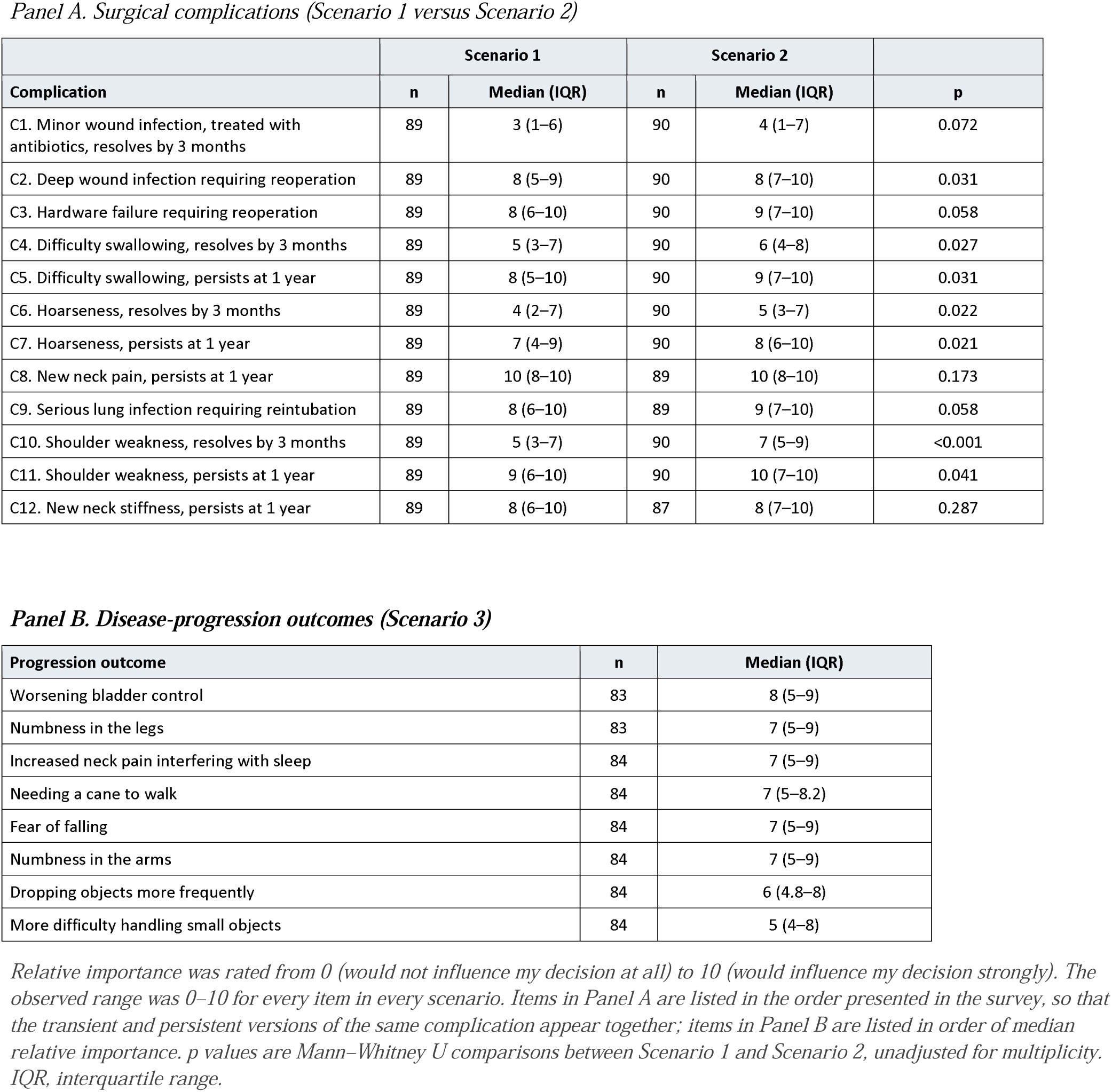
Relative importance of each adverse event, by randomized scenario.

|  | **Scenario 1** |  | **Scenario 2** |  |  |
|---|---|---|---|---|---|
| **Complication** | **n** | **Median (IQR)** | **n** | **Median (IQR)** | **p** |
| C1. Minor wound infection, treated with antibiotics, resolves by 3 months | 89 | 3 (1–6) | 90 | 4 (1–7) | 0.072 |
| C2. Deep wound infection requiring reoperation | 89 | 8 (5–9) | 90 | 8 (7–10) | 0.031 |
| C3. Hardware failure requiring reoperation | 89 | 8 (6–10) | 90 | 9 (7–10) | 0.058 |
| C4. Difficulty swallowing, resolves by 3 months | 89 | 5 (3–7) | 90 | 6 (4–8) | 0.027 |
| C5. Difficulty swallowing, persists at 1 year | 89 | 8 (5–10) | 90 | 9 (7–10) | 0.031 |
| C6. Hoarseness, resolves by 3 months | 89 | 4 (2–7) | 90 | 5 (3–7) | 0.022 |
| C7. Hoarseness, persists at 1 year | 89 | 7 (4–9) | 90 | 8 (6–10) | 0.021 |
| C8. New neck pain, persists at 1 year | 89 | 10 (8–10) | 89 | 10 (8–10) | 0.173 |
| C9. Serious lung infection requiring reintubation | 89 | 8 (6–10) | 89 | 9 (7–10) | 0.058 |
| C10. Shoulder weakness, resolves by 3 months | 89 | 5 (3–7) | 90 | 7 (5–9) | <0.001 |
| C11. Shoulder weakness, persists at 1 year | 89 | 9 (6–10) | 90 | 10 (7–10) | 0.041 |
| C12. New neck stiffness, persists at 1 year | 89 | 8 (6–10) | 87 | 8 (7–10) | 0.287 |
| **Progression outcome** | **n** | **Median (IQR)** |  |  |  |
| Worsening bladder control | 83 | 8 (5–9) |  |  |  |
| Numbness in the legs | 83 | 7 (5–9) |  |  |  |
| Increased neck pain interfering with sleep | 84 | 7 (5–9) |  |  |  |
| Needing a cane to walk | 84 | 7 (5–8.2) |  |  |  |
| Fear of falling | 84 | 7 (5–9) |  |  |  |
| Numbness in the arms | 84 | 7 (5–9) |  |  |  |
| Dropping objects more frequently | 84 | 6 (4.8–8) |  |  |  |
| More difficulty handling small objects | 84 | 5 (4–8) |  |  |  |

*Relative importance was rated from 0 (would not influence my decision at all) to 10 (would influence my decision strongly). The observed range was 0–10 for every item in every scenario. Items in Panel A are listed in the order presented in the survey, so that the transient and persistent versions of the same complication appear together; items in Panel B are listed in order of median relative importance. p values are Mann–Whitney U comparisons between Scenario 1 and Scenario 2, unadjusted for multiplicity. IQR, interquartile range.*

**Supplementary Table S2.** Treatment choice responses for each adverse event, by randomized scenario.

| Complication | Arm | n | Yes n (%) | Unsure n (%) | No n (%) |
|---|---|---|---|---|---|
| C1. Minor wound infection, treated with antibiotics, resolves by 3 months | S1 | 89 | 59 (66.3) | 17 (19.1) | 13 (14.6) |
|  | S2 | 89 | 48 (53.9) | 27 (30.3) | 14 (15.7) |
| C2. Deep wound infection requiring reoperation | S1 | 86 | 16 (18.6) | 31 (36.0) | 39 (45.3) |
|  | S2 | 89 | 12 (13.5) | 25 (28.1) | 52 (58.4) |
| C3. Hardware failure requiring reoperation | S1 | 89 | 14 (15.7) | 24 (27.0) | 51 (57.3) |
|  | S2 | 90 | 11 (12.2) | 20 (22.2) | 59 (65.6) |
| C4. Difficulty swallowing, resolves by 3 months | S1 | 89 | 45 (50.6) | 18 (20.2) | 26 (29.2) |
|  | S2 | 90 | 44 (48.9) | 27 (30.0) | 19 (21.1) |
| C5. Difficulty swallowing, persists at 1 year | S1 | 88 | 9 (10.2) | 24 (27.3) | 55 (62.5) |
|  | S2 | 89 | 9 (10.1) | 15 (16.9) | 65 (73.0) |
| C6. Hoarseness, resolves by 3 months | S1 | 89 | 56 (62.9) | 21 (23.6) | 12 (13.5) |
|  | S2 | 90 | 43 (47.8) | 32 (35.6) | 15 (16.7) |
| C7. Hoarseness, persists at 1 year | S1 | 88 | 24 (27.3) | 24 (27.3) | 40 (45.5) |
|  | S2 | 90 | 19 (21.1) | 24 (26.7) | 47 (52.2) |
| C8. New neck pain, persists at 1 year | S1 | 89 | 7 (7.9) | 13 (14.6) | 69 (77.5) |
|  | S2 | 90 | 7 (7.8) | 4 (4.4) | 79 (87.8) |
| C9. Serious lung infection requiring reintubation | S1 | 89 | 11 (12.4) | 31 (34.8) | 47 (52.8) |
|  | S2 | 90 | 13 (14.4) | 21 (23.3) | 56 (62.2) |
| C10. Shoulder weakness, resolves by 3 months | S1 | 88 | 46 (52.3) | 26 (29.5) | 16 (18.2) |
|  | S2 | 90 | 40 (44.4) | 31 (34.4) | 19 (21.1) |
| C11. Shoulder weakness, persists at 1 year | S1 | 89 | 10 (11.2) | 20 (22.5) | 59 (66.3) |
|  | S2 | 90 | 10 (11.1) | 11 (12.2) | 69 (76.7) |
| C12. New neck stiffness, persists at 1 year | S1 | 89 | 11 (12.4) | 28 (31.5) | 50 (56.2) |
|  | S2 | 90 | 13 (14.4) | 23 (25.6) | 54 (60.0) |
| **Progression outcome** | **n** | **Yes n (%) [95% CI]** | **Unsure n (%) [95% CI]** | **No n (%) [95% CI]** |  |
| Worsening bladder control | 83 | 14 (16.9) [10.3–26.3] | 22 (26.5) [18.2–36.9] | 47 (56.6) [45.9–66.8] |  |
| Increased neck pain interfering with sleep | 83 | 13 (15.7) [9.4–25.0] | 26 (31.3) [22.4–41.9] | 44 (53.0) [42.4–63.4] |  |
| Numbness in the legs | 83 | 14 (16.9) [10.3–26.3] | 27 (32.5) [23.4–43.2] | 42 (50.6) [40.1–61.1] |  |
| Numbness in the arms | 84 | 17 (20.2) [13.0–30.0] | 27 (32.1) [23.1–42.7] | 40 (47.6) [37.3–58.2] |  |
| Fear of falling | 84 | 21 (25.0) [17.0–35.2] | 26 (31.0) [22.1–41.5] | 37 (44.0) [33.9–54.7] |  |
| Dropping objects more frequently | 83 | 25 (30.1) [21.3–40.7] | 25 (30.1) [21.3–40.7] | 33 (39.8) [29.9–50.5] |  |
| Needing a cane to walk | 84 | 21 (25.0) [17.0–35.2] | 36 (42.9) [32.8–53.5] | 27 (32.1) [23.1–42.7] |  |
| More difficulty handling small objects | 84 | 28 (33.3) [24.2–43.9] | 29 (34.5) [25.2–45.2] | 27 (32.1) [23.1–42.7] |  |

*In Panel A, “yes” indicates the respondent would still choose surgery and “no” that they would decline it; in Panel B, “yes” indicates they would continue nonoperative treatment and “no” that they would elect surgery. The “no” column is the dominant decision factor rate reported in Table 2. Denominators differ slightly from Supplementary Table S1 for some items because the two questions were answered independently and a small number of respondents completed one but not the other. Confidence intervals are Wilson score intervals.*

**Supplementary Figure 1:**
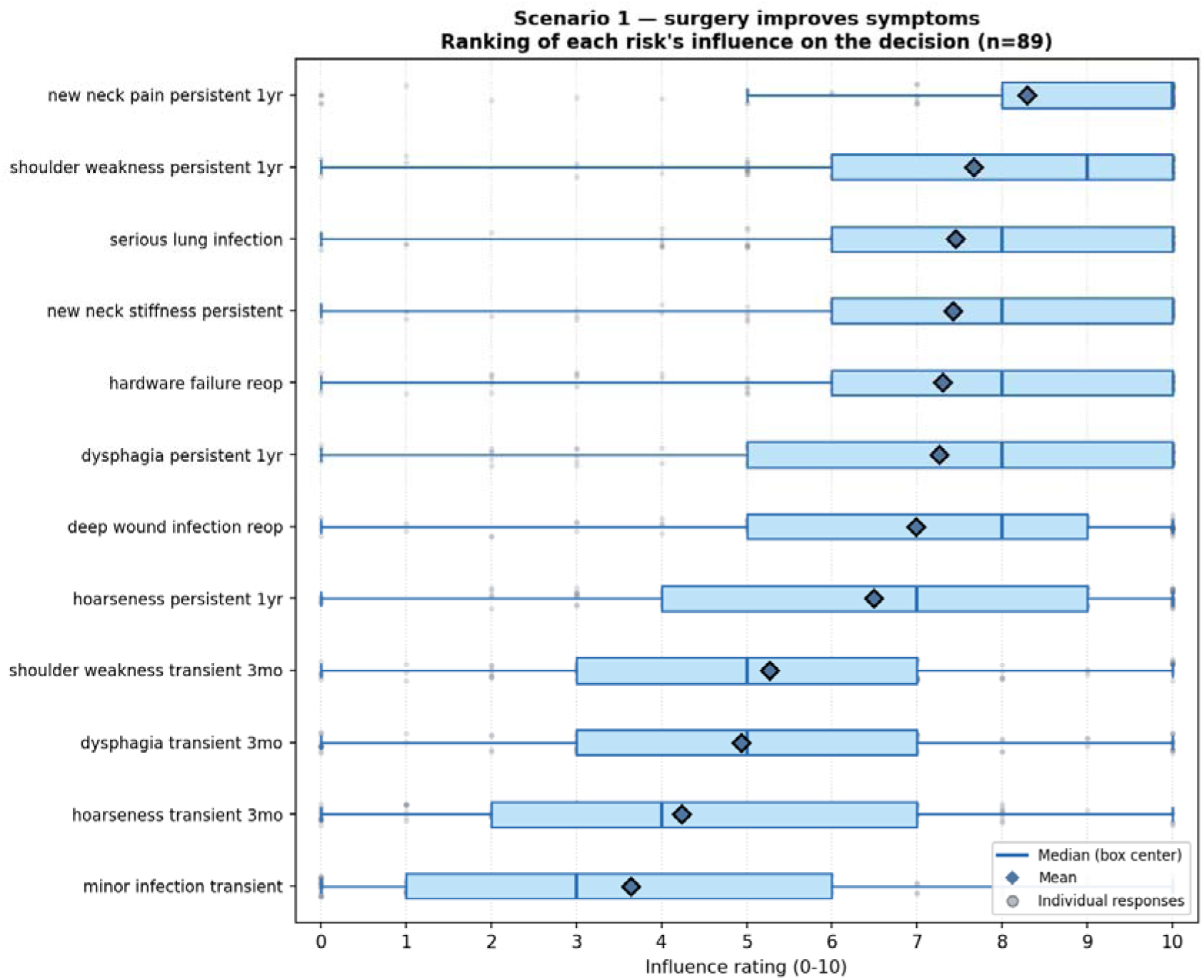
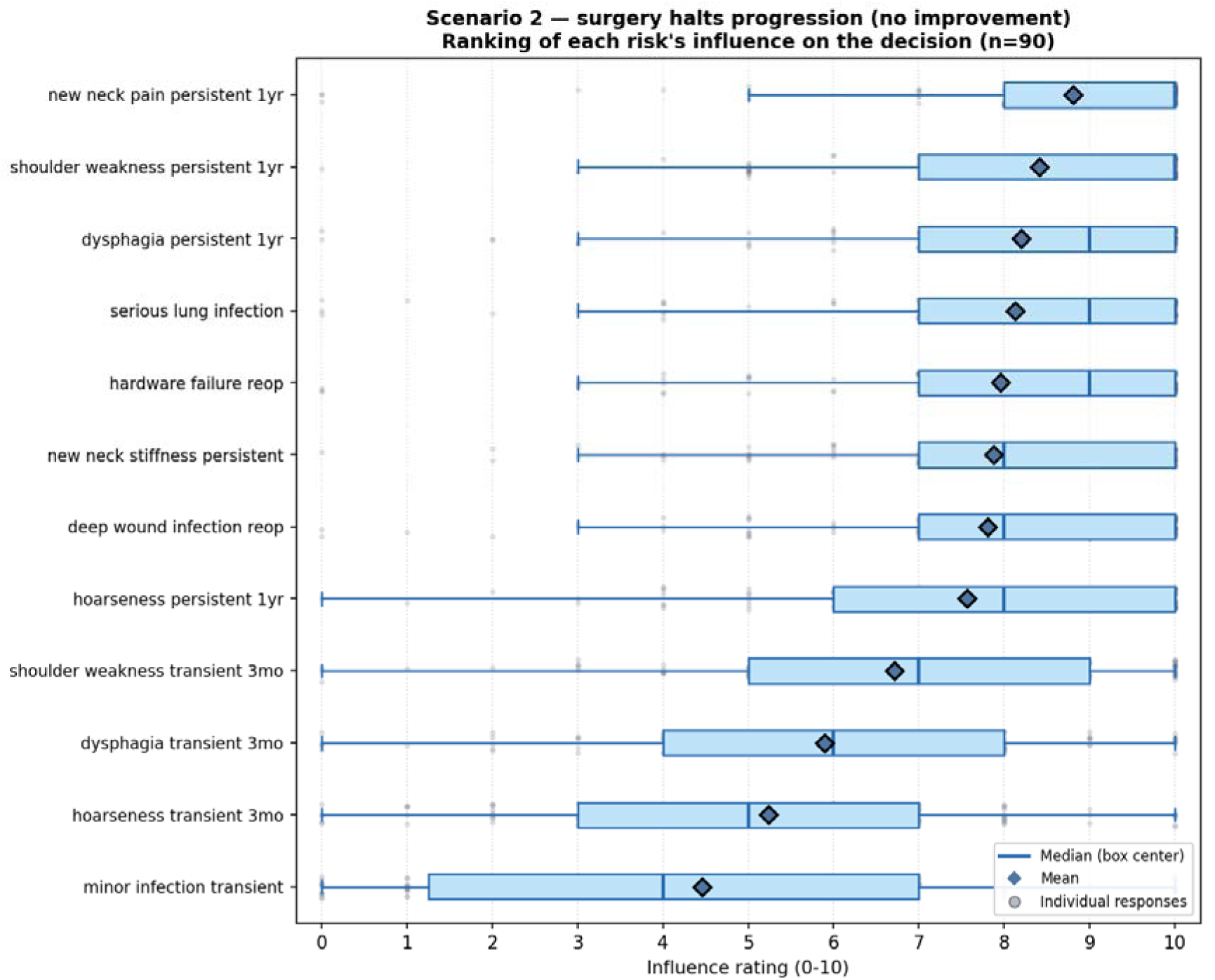
Influence of each complication on the decision about surgery, by benefit frame. (a) Scenario 1, surgery improves symptoms (n = 89). (b) Scenario 2, surgery halts progression (n = 90). Boxes show median and interquartile range of the 0–10 influence rating; diamonds show means; grey points show individual responses.

**Supplementary Figure 2:**
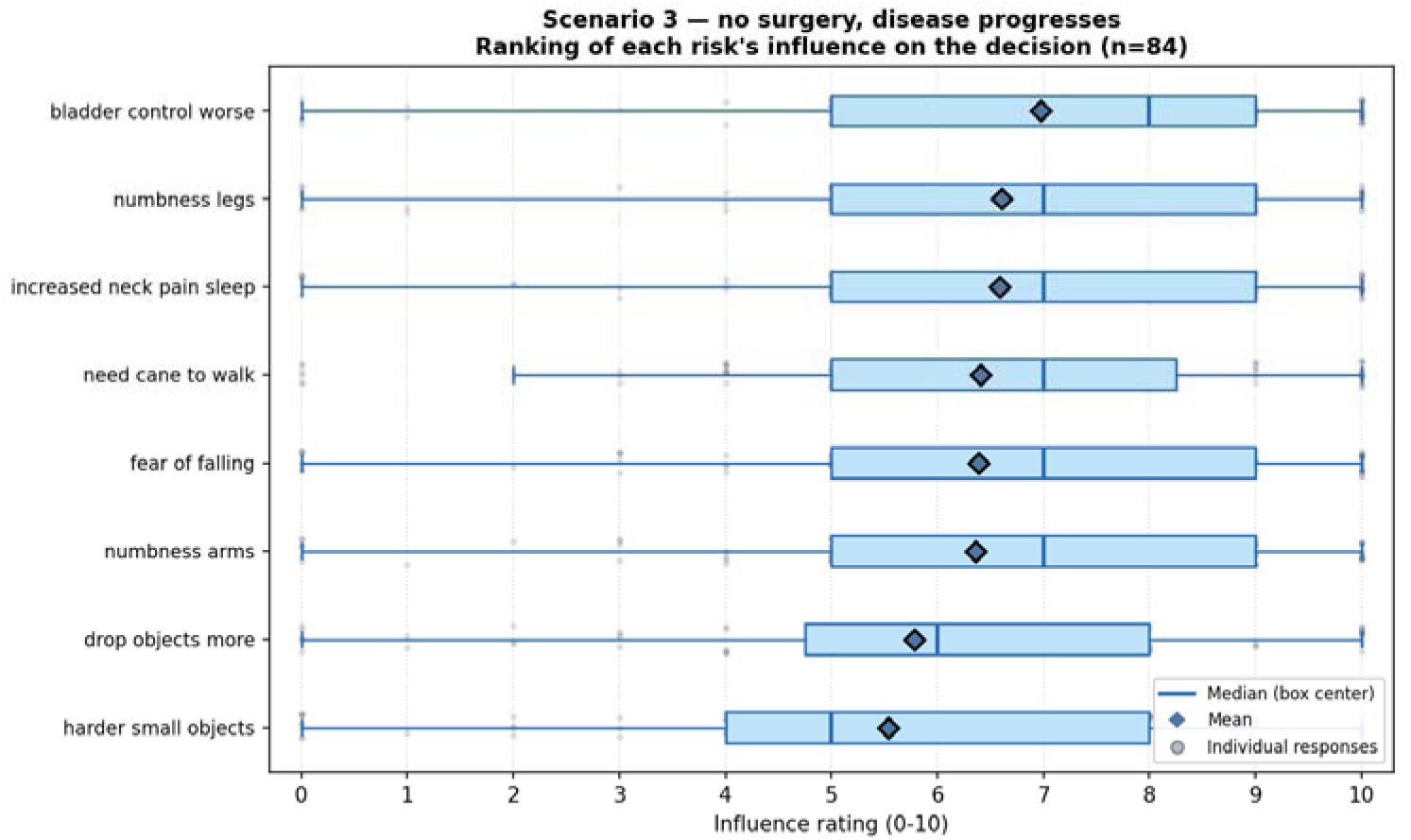
Influence of each disease-progression outcome on the decision in the observation arm (Scenario 3, n = 84). Boxes show median and interquartile range of the 0–10 influence rating; diamonds show means; grey points show individual responses.

## References

1. Badhiwala JH, Ahuja CS, Akbar MA, et al. Degenerative cervical myelopathy — update and future directions. Nat Rev Neurol. 2020;16(2):108–124. doi:10.1038/s41582-019-0303-0

2. Nouri A, Tetreault L, Singh A, Karadimas SK, Fehlings MG. Degenerative Cervical Myelopathy: Epidemiology, Genetics, and Pathogenesis. Spine (Phila Pa 1976). 2015;40(12):E675-E693. doi:10.1097/BRS.0000000000000913

3. Smith SS, Stewart ME, Davies BM, Kotter MRN. The Prevalence of Asymptomatic and Symptomatic Spinal Cord Compression on Magnetic Resonance Imaging: A Systematic Review and Meta-analysis. Global Spine J. 2021;11(4):597–607. doi:10.1177/2192568220934496

4. Pope DH, Mowforth OD, Davies BM, Kotter MRN. Diagnostic Delays Lead to Greater Disability in Degenerative Cervical Myelopathy and Represent a Health Inequality. Spine (Phila Pa 1976). 2020;45(6):368-377. doi:10.1097/BRS.0000000000003305

5. Davies BM, Mowforth OD, Smith EK, Kotter MRN. Degenerative cervical myelopathy. BMJ. 2018;360:k186. doi:10.1136/bmj.k186

6. Tetreault L, Kopjar B, Nouri A, et al. The modified Japanese Orthopaedic Association scale: establishing criteria for mild, moderate and severe impairment in patients with degenerative cervical myelopathy. Eur Spine J. 2017;26(1):78–84. doi:10.1007/s00586-016-4660-8

7. Karim SM, Cadotte DW, Wilson JR, et al. Effectiveness of Surgical Decompression in Patients With Degenerative Cervical Myelopathy: Results of the Canadian Prospective Multicenter Study. Neurosurgery. 2021;89(5):844–851. doi:10.1093/neuros/nyab295

8. Kögl N, Evaniew N, McIntosh G, et al. Outcome of Patients With Mild Degenerative Cervical Myelopathy Treated Nonoperatively: An Observational Study From the Canadian Spine Outcome and Research Network. Neurosurgery. 2026;99(1):60–67. doi:10.1227/neu.

9. Evaniew N, Cadotte DW, Dea N, et al. Deterioration After Surgery for Degenerative Cervical Myelopathy: An Observational Study From the Canadian Spine Outcomes and Research Network. Spine (Phila Pa 1976). 2023;48(5):310-320. doi:10.1097/BRS.0000000000004552

10. Fehlings MG, Tetreault LA, Riew KD, et al. A Clinical Practice Guideline for the Management of Patients With Degenerative Cervical Myelopathy. Global Spine J. 2017;7(3 Suppl):70S-83S. doi:10.1177/2192568217701914

11. Eysenbach G. Improving the Quality of Web Surveys: the Checklist for Reporting Results of Internet E-Surveys (CHERRIES). J Med Internet Res. 2004;6(3):e34. doi:10.2196/jmir.6.3.e34

12. Chandler J, Rosenzweig C, Moss AJ, Robinson J, Litman L. Online panels in social science research: Expanding sampling methods beyond Mechanical Turk. Behav Res Methods. 2019;51(5):2022–2038. doi:10.3758/s13428-019-01273-7

13. Greszki R, Meyer M, Schoen H. Exploring the effects of removing “too fast” responses and respondents from web surveys. Public Opin Q. 2015;79(2):471–503. doi:10.1093/poq/nfu058

14. Wilson EB. Probable Inference, the Law of Succession, and Statistical Inference. J Am Stat Assoc. 1927;22(158):209–212. doi:10.1080/01621459.1927.10502953

15. Newcombe RG. Interval estimation for the difference between independent proportions: comparison of eleven methods. Stat Med. 1998;17(8):873–890. doi:10.1002/(sici)1097-0258(19980430)17:8<873::aid-sim779>3.0.co;2-i

17. Yakdan S, Joseph K, Zhang J, et al. Prevalence of Sleep Disturbance and Associated Risk Factors in Degenerative Cervical Myelopathy. Journal of Clinical Medicine. 2025; 14(19):7110. 10.3390/jcm14197110

18. Ghogawala Z, Terrin N, Dunbar MR, et al. Effect of Ventral vs Dorsal Spinal Surgery on Patient-Reported Physical Functioning in Patients With Cervical Spondylotic Myelopathy: A Randomized Clinical Trial. JAMA. 2021;325(10):942–951. doi:10.1001/jama.2021.1233

19. Yakdan S, Joseph K, Poulin N, et al. Disparities in quality of life and health literacy among patients with degenerative cervical myelopathy: the influence of racial and ethnic factors in the All of Us Research Program. SPI. 2026;44(6):920–929. doi:10.3171/2025.11.SPINE25662

20. Arkam F, Zeng X, Goldstein E, et al. Patient and Surgeon Willingness to Participate in a Randomized Trial of Surgery Versus Observation for Mild Cervical Spondylotic Myelopathy: A Cross-Sectional Survey Study. medRxiv. Preprint posted online August 18, 2026. doi:10.64898/2026.08.18.26360719

21. Singh P, Gonuguntla S, Chen E, et al. A Qualitative Study of Patient and Healthcare Provider Perspectives on Mobile Health Assessments for Cervical Spondylotic Myelopathy. medRxiv. Preprint posted online March 5, 2026. doi:10.64898/2026.03.04.26347622

