## Supplementary Material - Survey for "Tolerance for Adverse Events from Operative and Nonoperative Treatment for Mild Cervical Spondylotic Myelopathy"

### Survey: Decision-Making in Early Stage Compression of the Spinal Cord in the Neck

You are invited to participate in a research study being led by investigators from Washington University in St. Louis.

**What is the goal of this study?**

The goal of this study is to understand how people feel about treatment for early-stage compression of the spinal cord in the neck. This type of compression typically results from age-related arthritis. Symptoms from this disease can cause numbness in the hands or feet, hand clumsiness, unsteadiness with walking, or increased urgency to use the bathroom. We are interested in studying whether surgery is a beneficial treatment for early-stage disease.

**What will I do during the study?**

You will view different information about early-stage compression of the spine. We will ask you to imagine that you are going through a scenario as a patient, and we will ask you how you would feel about different potential outcomes.

You can take as much time as you need to review the page. Then you will be asked questions about what you learned and how you felt about the imagined scenario.

This survey should take no more than 10 minutes to complete.

**What are the risks of this study?**

There are no risks of being in this study.

**What are the benefits of this study?**

We do not expect you to benefit from this study. What we learn might help other clinicians, patients, and researchers in the future.

**Will I be compensated?**

Upon completion of the study, you will receive compensation in the amount you have agreed to with the platform through which you entered this survey.

**Do I have to participate in this study?**

No, you do not have to participate in this study. It is your choice, and you can stop at any time.

If you have any questions about the research study, please contact Jacob Greenberg

If you have questions, concerns, or complaints about your rights as a research participant, please contact the Human Research Protection Office at 1-(800)-438-0445 or. Thank you very much for your consideration of this research study.

#### Section 1: Consent

Do you agree to participate in this anonymous survey?

☐ Yes

☐ No

#### Section 2: Participant Characteristics

1) Sex at birth

☐ Male

☐ Female

☐ Other

1a) What gender do you identify with?

☐ Male

☐ Female

☐ Non-binary

☐ Prefer to self-describe: ___________

2) What is your age?

☐ 40–49

☐ 50–59

☐ 60–69

☐ 70–79

☐ 80+

☐ Prefer not to say

3) What is the highest level of education you have completed?

☐ Less than a high school degree

☐ High school degree or GED

☐ Some college or Associate’s Degree

☐ Bachelor’s degree

☐ Graduate or professional degree

☐ Prefer not to say

4) What is your current marital status?

☐ Single / Never married

☐ Married or living with a partner

☐ Divorced

☐ Widowed

☐ Prefer not to say

5) What type of health insurance do you currently have?

☐ Private insurance (through employer or individual plan)

☐ Medicare

☐ Medicaid

☐ Military / Veterans (VA, TRICARE)

☐ Other public insurance

☐ Uninsured

☐ Prefer not to say

6) What is your current employment status?

☐ Currently employed

☐ Retired

☐ Homemaker

☐ Unemployed

☐ On disability

☐ Prefer not to say

7) Have you ever had major surgery before? ☐ No ☐ Yes

#### Section 3: Hypothetical Scenario

Now we would like you to imagine the following situation:

You have pressure on the spinal cord in your neck from arthritis that has caused narrowing in your spine. This type of pressure on the spinal cord might affect walking, balance, the ability to use your hands (e.g., clumsiness), and sometimes your ability to control your bladder. Pain can also be a symptom of this disease.

Your condition is mild. This means you have a little trouble working with small items. You take more time to button a shirt or use a zipper, and opening jars has become more difficult, but you can still do it. Your balance is slightly off. You can walk on flat ground without problems, but you avoid walking on uneven or slippery surfaces because you are slightly unsteady. You can otherwise take care of yourself without help from others.

As arthritis progresses, this condition can worsen over time, but it tends to be slow, typically worsening over months to years. Some people with mild symptoms choose to have surgery to relieve pressure on the spinal cord and keep their symptoms from getting worse. They also might see some improvement in these symptoms. Others choose to keep track of their symptoms, knowing they may remain unchanged for years, because they are ok living with these mild symptoms, and they worry about complications from surgery. There is no one-size-fits-all answer on how to manage mild symptoms from spinal cord pressure and arthritis.

#### Instructions

In the following sections, you will read a scenario. For each outcome, rate how strongly it would influence your decision to pursue surgery.

Use the following scale:
0 = Would NOT influence my decision at all
10 = Would influence my decision strongly

#### Scenario 1: Surgery improves your symptoms

You choose to have surgery to relieve some symptoms and prevent further worsening of your symptoms. **One year later**, your symptoms (for example, your balance, hand function, and numbness) are **overall** **improved** and are now less bothersome. However, you have a complication from surgery. How strongly would the *risk* of each complication influence your decision about whether or not to have surgery

1.
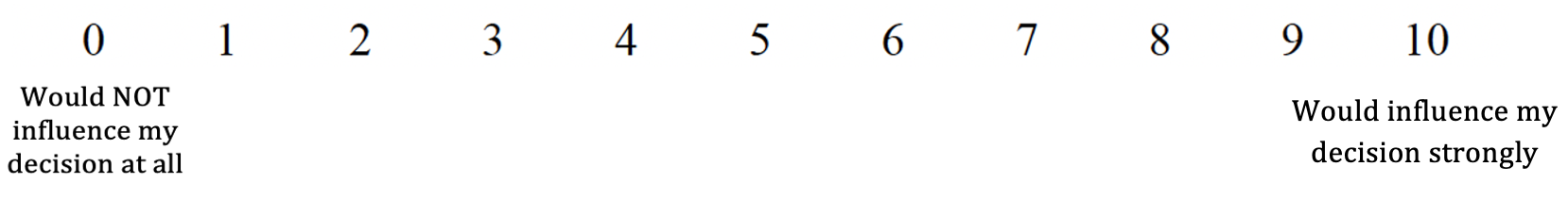
After surgery, you get a minor infection that needs to be treated by taking pills by mouth for 1-2 weeks. You do not need to stay in the hospital. How important would this complication be to your decision about whether or not to have surgery?

Would you still choose to have surgery, even if you had this complication?

☐Yes

☐No

☐Unsure

2.
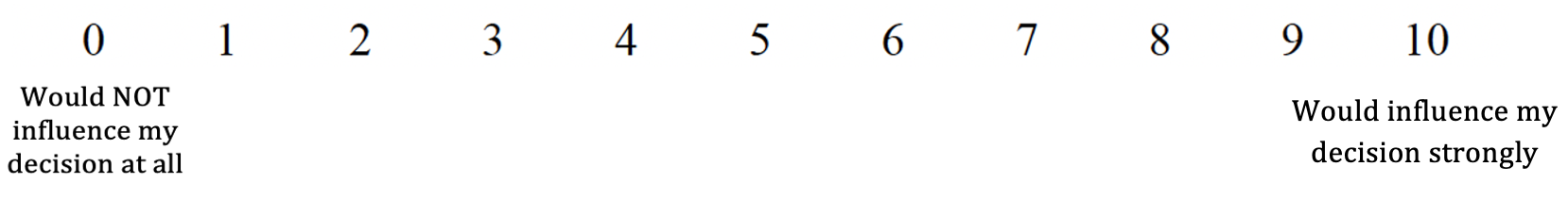
After surgery, you get an infection deep inside the wound that needs more surgery to treat it. You have to stay in the hospital for another week and then have to inject antibiotics into your blood at home for another month afterwards. How important would this complication be to your decision about whether or not to have surgery?

Would you still choose to have surgery, even if you had this complication?

☐Yes

☐No

☐Unsure

3.
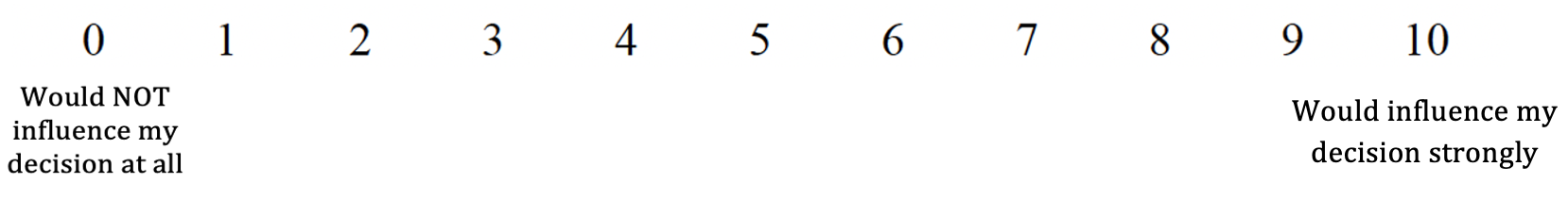
After surgery, a problem occurs with the rods and screws used during your surgery to remove pressure on the spine, and you need to have another surgery to fix it. How important would this complication be to your decision about whether or not to have surgery?

Would you still choose to have surgery, even if you had this complication?

☐Yes

☐No

☐Unsure

4.
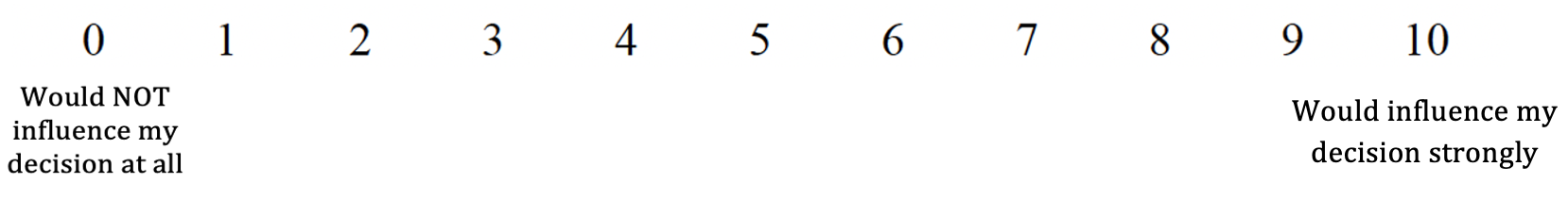
After surgery, you start having difficulty swallowing that makes you eat slowly, use extra effort for solid food, and avoid certain thicker foods. After 3 months, your eating returns to normal. How important would this complication be to your decision about whether or not to have surgery?

Would you still choose to have surgery, even if you had this complication?

☐Yes

☐No

☐Unsure

5. After surgery, you develop difficulty swallowing that makes eat slowly, use extra effort for solid food, and avoid certain thicker foods. 1 year after surgery, you still have this problem. How important would this complication be to your decision about whether or not to have surgery?

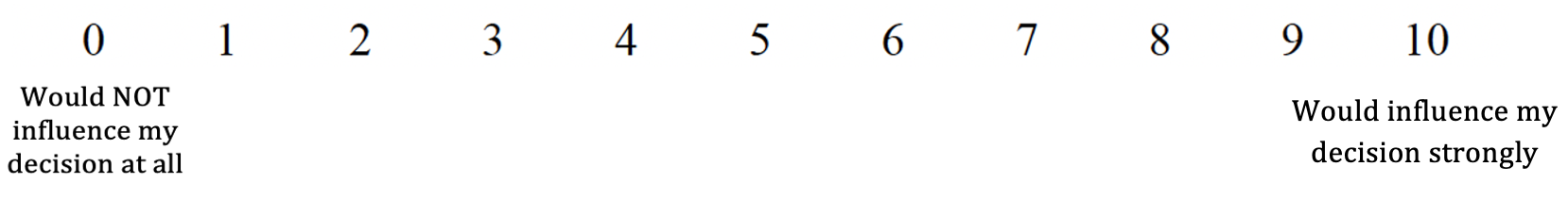

Would you still choose to have surgery, even if you had this complication?

☐ Yes

☐ No

☐ Unsure

6.
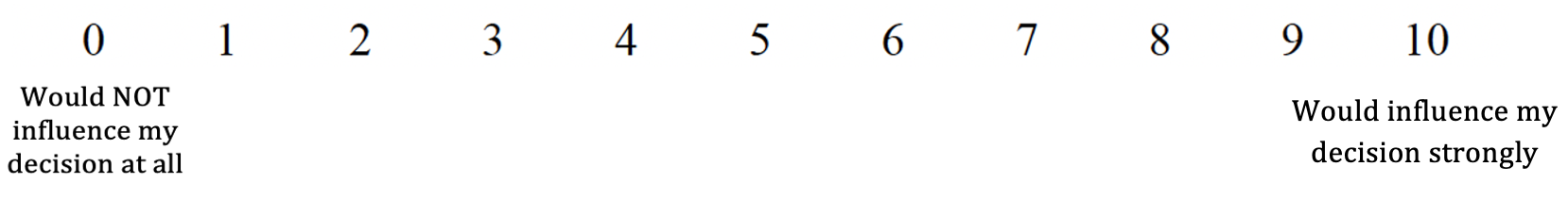
After surgery, your voice becomes hoarse. You can still speak, but it is harder for others to hear as well. After 3 months, your voice returns to normal**.** How important would this complication be to your decision about whether or not to have surgery?

Would you still choose to have surgery, even if you had this complication?

☐Yes

☐No

☐Unsure

7.
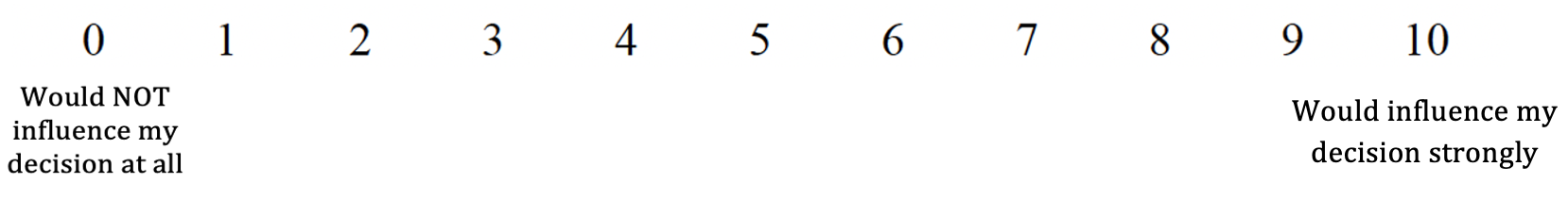
 After surgery, your voice becomes hoarse. You can still speak, but it is harder for others to hear as well. 1 year after surgery, you still have this problem. How important would this complication be to your decision about whether or not to have surgery?

Would you still choose to have surgery, even if you had this complication?

☐ Yes

☐ No

☐ Unsure

8.
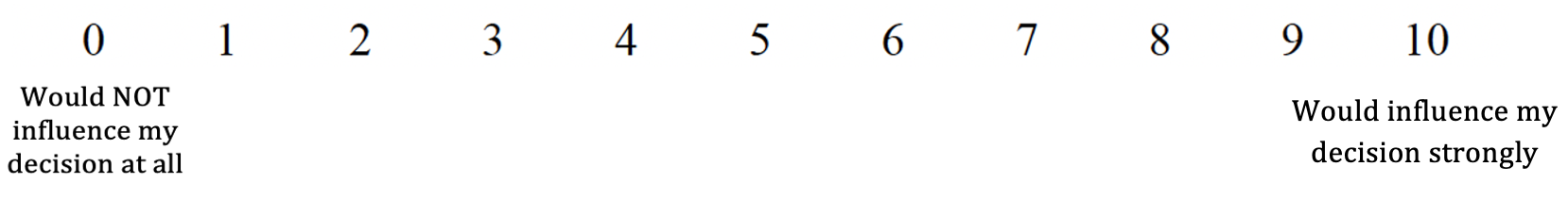
After surgery, you develop *new* neck pain that is worse than it was before surgery. 1 year after surgery, this pain continues to impact your day-to-day activities and the quality of your sleep. How important would this complication be to your decision about whether or not to have surgery?

Would you still choose to have surgery, even if you had this complication?

☐ Yes

☐ No

☐ Unsure

9.
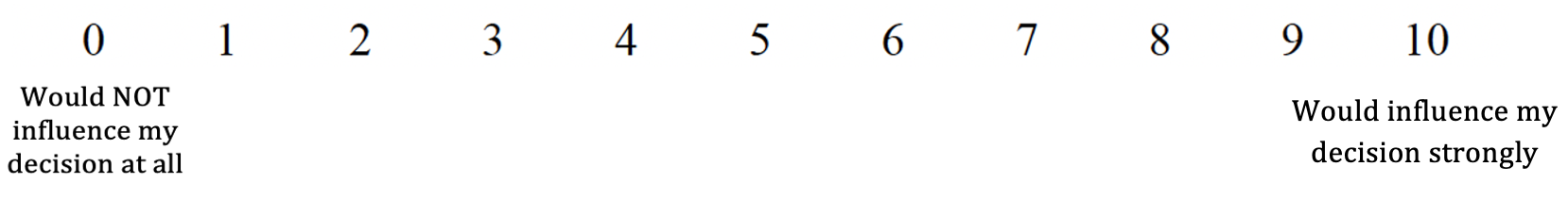
After surgery, you get a serious lung infection. You need to have the breathing tube put back in for a week and spend a total of a month in the hospital. 3 months later, you make a full recovery. How important would this complication be to your decision about whether or not to have surgery?

Would you still choose to have surgery, even if you had this complication?

☐Yes

☐No

☐Unsure

10.
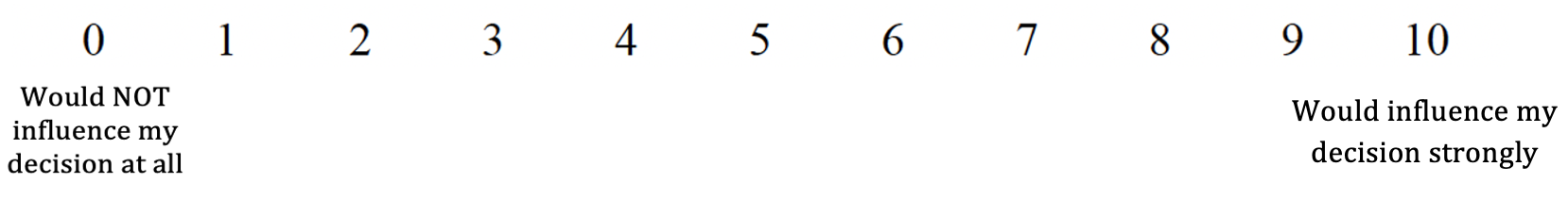
After surgery, you get weak in one shoulder that makes it hard to lift that arm up in the air, making it hard to use that arm for things like brushing your teeth. After 3 months, your shoulder function returns to normal. How important would this complication be to your decision about whether or not to have surgery?

Would you still choose to have surgery, even if you had this complication?

☐Yes

☐No

☐Unsure

11.
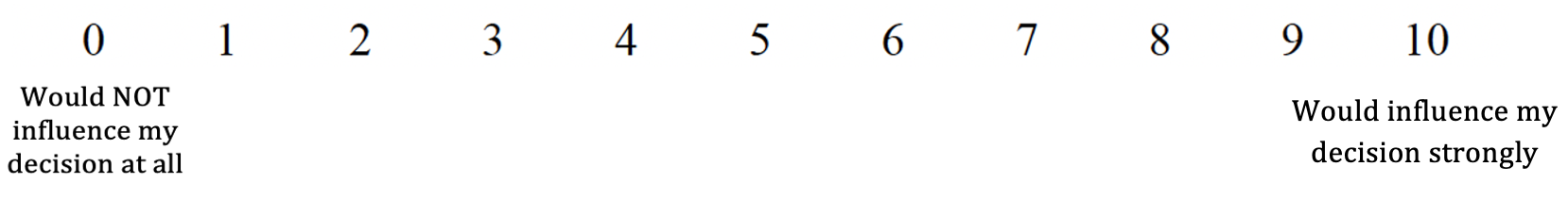
 After surgery, you get weak in one shoulder that makes it hard to lift that arm up in the air, making it hard to use that arm for things like brushing your teeth. 1 year after surgery, your shoulder function is still no better. How important would this complication be to your decision about whether or not to have surgery?

Would you still choose to have surgery, even if you had this complication?

☐ Yes

☐ No

☐ Unsure

12.
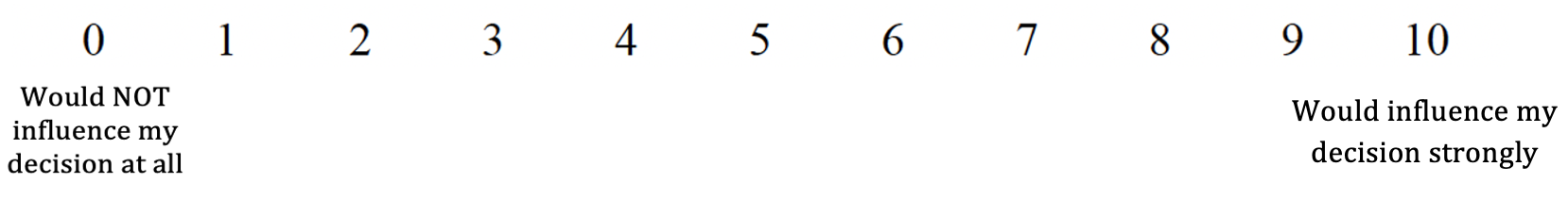
After surgery, you develop new stiffness in your neck. You can still drive, but it’s harder. The decreased mobility is noticeable in your everyday activities**.** How important would this complication be to your decision about whether or not to have surgery?

Would you still choose to have surgery, even if you had this complication?

☐ Yes

☐ No

☐ Unsure

Which of the following is typically cold?

☐ Fire

☐ Ice

☐ Boiling water

☐ Sunlight

#### Scenario 2: Surgery halts progression (no improvement)

You choose surgery. After recovery, your symptoms (for example, your balance, hand function, and numbness) are **stable**. Meaning you do **not** feel better overall, but your spinal cord compression is now gone, and your symptoms are **not** worsening. In this situation, how strongly would the *risk* of each complication influence your decision about whether or not to have surgery?

1. After surgery, you get a minor infection that needs to be treated by taking pills by mouth for 1-2 weeks. You do not need to stay in the hospital. How important would this complication be to your decision about whether or not to have surgery?

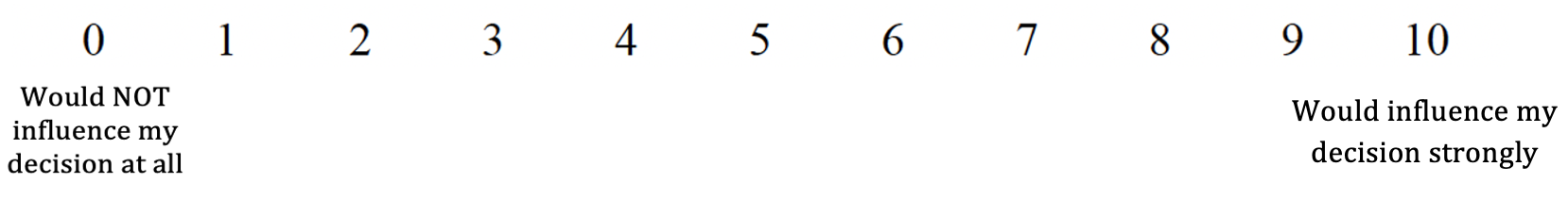

Would you still choose to have surgery, even if you had this complication?

☐Yes

☐No

☐Unsure

2.
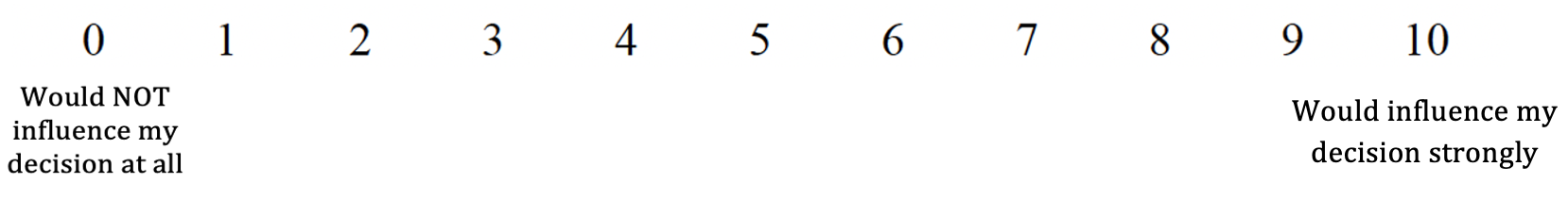
After surgery, you get an infection deep inside the wound that needs more surgery to treat it. You have to stay in the hospital for another week and then have to inject antibiotics into your blood at home for another month afterwards. How important would this complication be to your decision about whether or not to have surgery?

Would you still choose to have surgery, even if you had this complication?

☐Yes

☐No

☐Unsure

3.
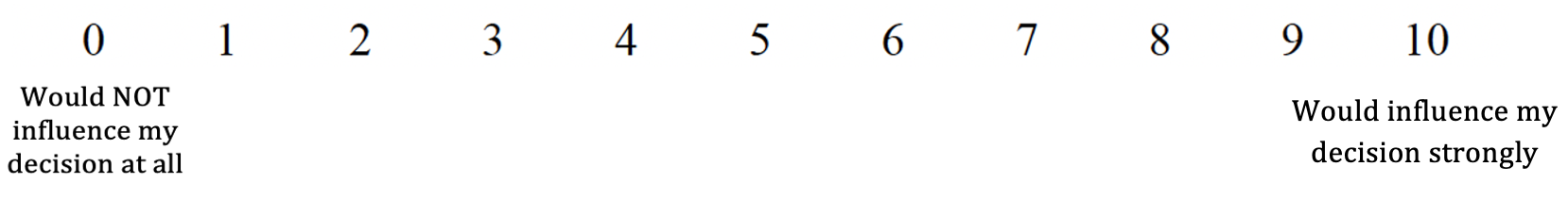
After surgery, a problem occurs with the rods and screws used during your surgery to remove pressure on the spine, and you need to have another surgery to fix it. How important would this complication be to your decision about whether or not to have surgery?

Would you still choose to have surgery, even if you had this complication?

☐Yes

☐No

☐Unsure

4. After surgery, you start having difficulty swallowing that makes you eat slowly, use extra effort for solid food, and avoid certain thicker foods. After 3 months, your eating returns to normal. How important would this complication be to your decision about whether or not to have surgery?

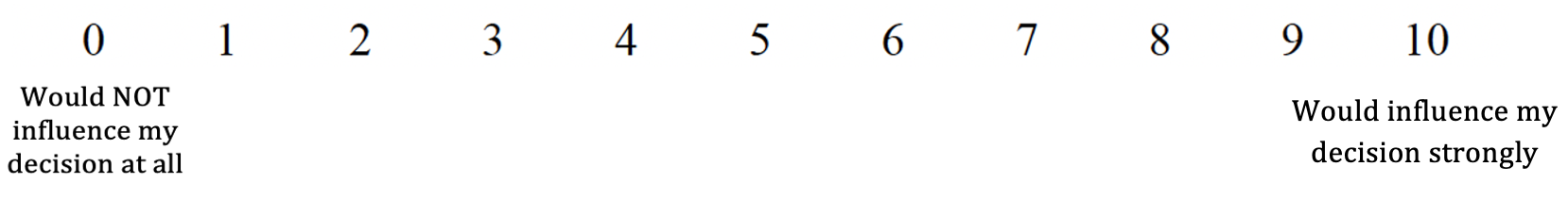

Would you still choose to have surgery, even if you had this complication?

☐Yes

☐No

☐Unsure

5.
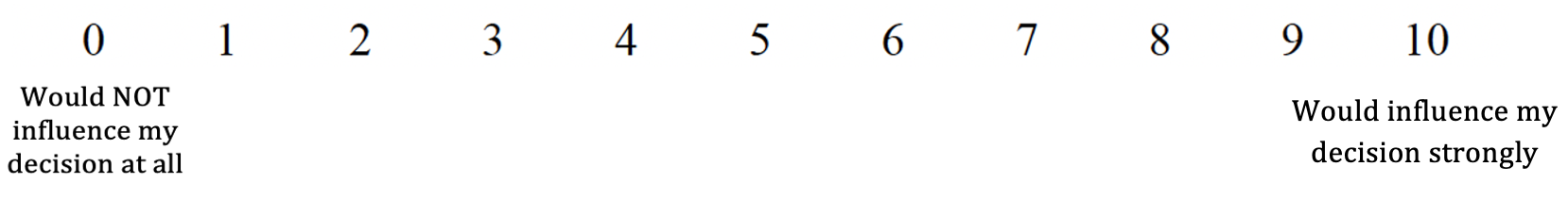
After surgery, you develop difficulty swallowing that makes eat slowly, use extra effort for solid food, and avoid certain thicker foods. 1 year after surgery, you still have this problem. How important would this complication be to your decision about whether or not to have surgery?

Would you still choose to have surgery, even if you had this complication?

☐ Yes

☐ No

☐ Unsure

6.
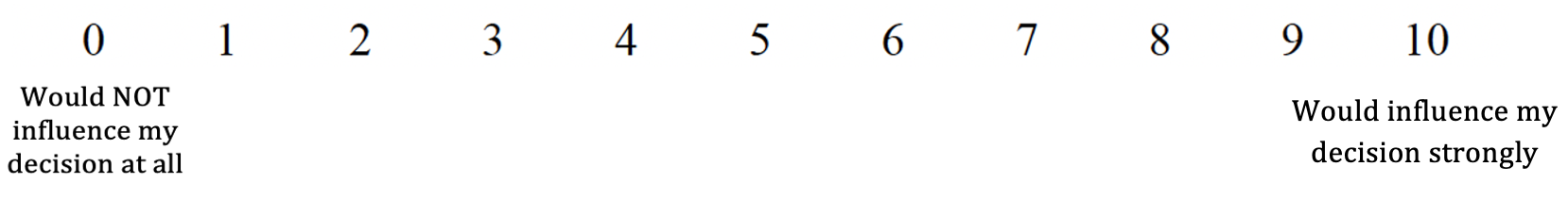
After surgery, your voice becomes hoarse. You can still speak, but it is harder for others to hear as well. After 3 months, your voice returns to normal. How important would this complication be to your decision about whether or not to have surgery?

Would you still choose to have surgery, even if you had this complication?

☐Yes

☐No

☐Unsure

7. After surgery, your voice becomes hoarse. You can still speak, but it is harder for others to hear as well. 1 year after surgery, you still have this problem. How important would this complication be to your decision about whether or not to have
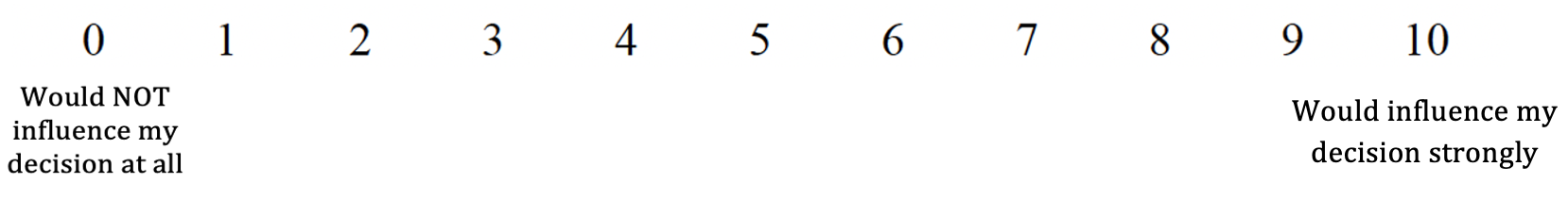
surgery?

Would you still choose to have surgery, even if you had this complication?

☐ Yes

☐ No

☐ Unsure

8.
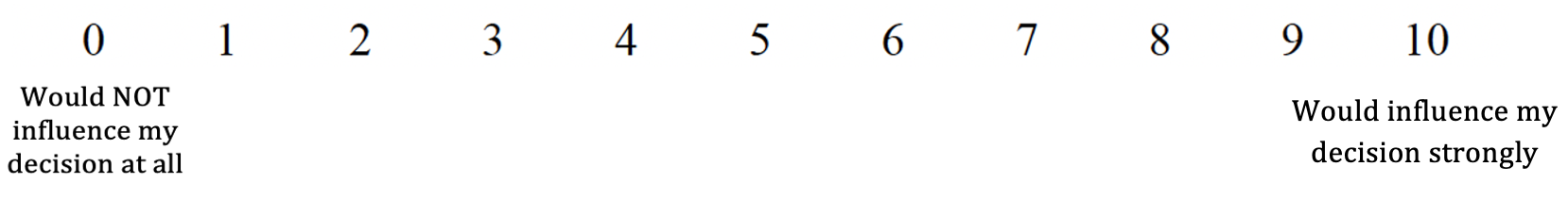
After surgery, you develop *new* neck pain that is worse than it was before surgery. 1 year after surgery**,** this pain continues to impact your day-to-day activities and the quality of your sleep. How important would this complication be to your decision about whether or not to have surgery?

Would you still choose to have surgery, even if you had this complication?

☐ Yes

☐ No

☐ Unsure

9. After surgery, you get a serious lung infection. You need to have the breathing tube put back in for a week and spend a total of a month in the hospital. 3 months later, you make a full recovery. How important would this complication be to your decision about whether or not to have surgery?

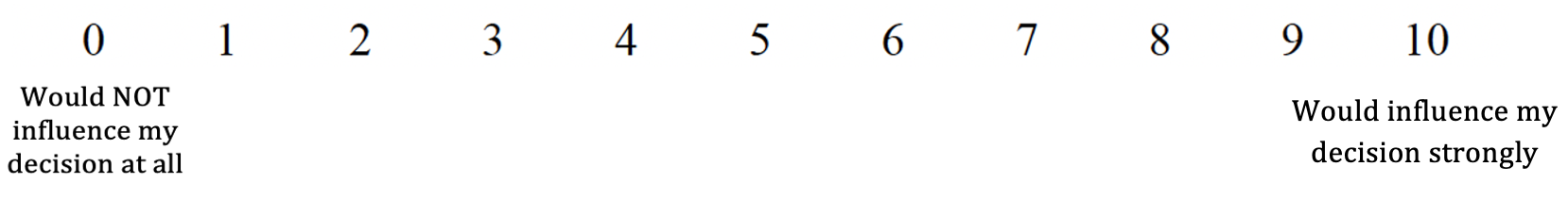

Would you still choose to have surgery, even if you had this complication?

☐Yes

☐No

☐Unsure

10. After surgery, you get weak in one shoulder that makes it hard to lift that arm up in the air, making it hard to use that arm for things like brushing your teeth. After 3 months, your shoulder function returns to normal. How important would this
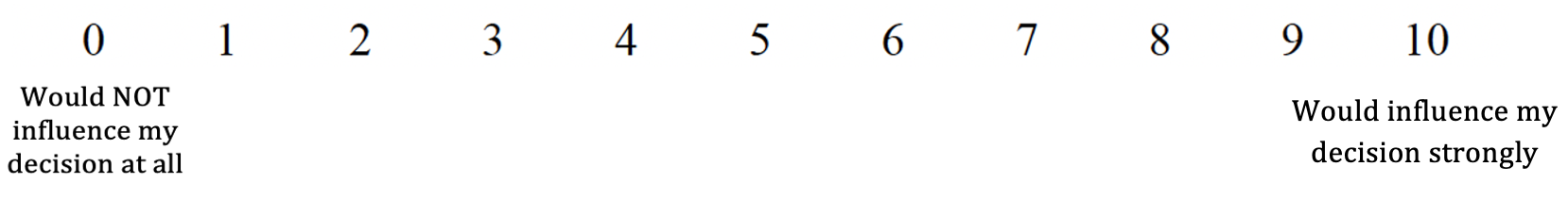
complication be to your decision about whether or not to have surgery?

Would you still choose to have surgery, even if you had this complication?

☐Yes

☐No

☐Unsure

11.
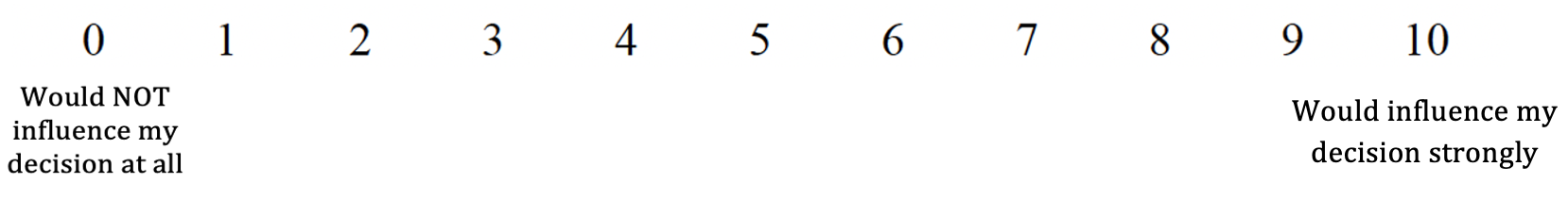
 After surgery, you get weak in one shoulder that makes it hard to lift that arm up in the air, making it hard to use that arm for things like brushing your teeth. 1 year after surgery, your shoulder function is still no better. How important would this complication be to your decision about whether or not to have surgery?

Would you still choose to have surgery, even if you had this complication?

☐ Yes

☐ No

☐ Unsure

12.
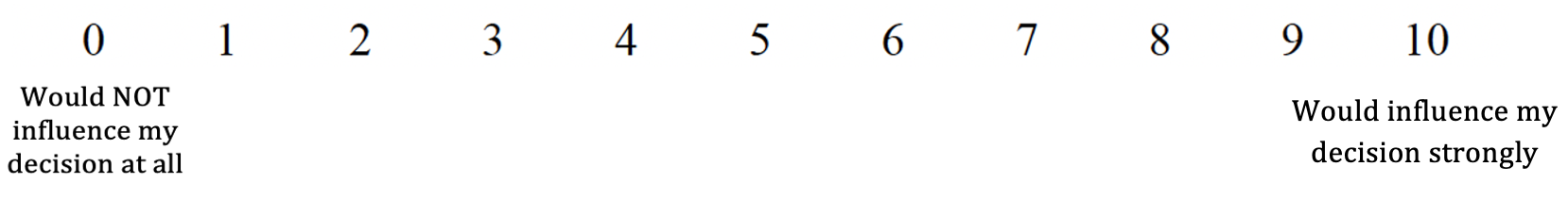
After surgery, you develop new stiffness in your neck. You can still drive, but it’s harder. The decreased mobility is noticeable in your everyday activities**.** How important would this complication be to your decision about whether or not to have surgery?

Would you still choose to have surgery, even if you had this complication?

☐ Yes

☐ No

☐ Unsure

Which of the following is typically cold?

☐ Fire

☐ Ice

☐ Boiling water

☐ Sunlight

#### Scenario 3: No surgery and disease progresses

You choose **not** to have surgery and instead monitor your symptoms. Over time, your spinal cord compression worsens, and new problems develop. Some of them could be permanent. If you had information about the risk of developing new problems like the ones below, how much would that influence your decision to choose to monitor your symptoms without surgery? Please answer each question thinking about each new problem described.

1. You now need a cane to walk safely. One year earlier, you did not need a cane and only had mild unsteadiness. How much would this outcome influence your decision about whether or not to have surgery?

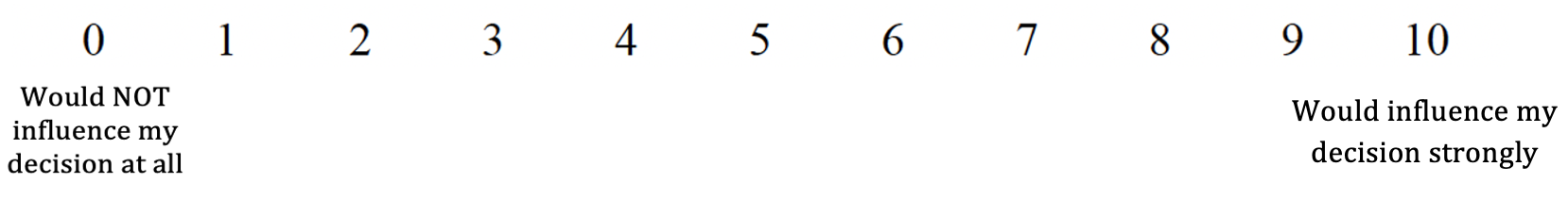

Would you still choose **not** to have surgery, even if you had this outcome?

☐Yes

☐No

☐Unsure

2.
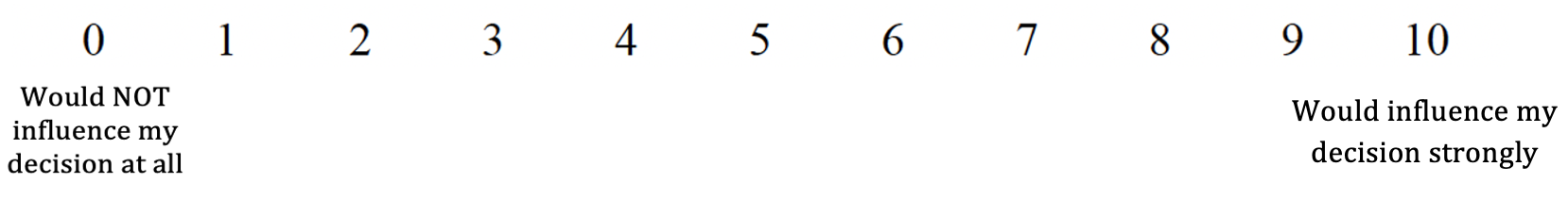
You have a new fear of falling because your unsteadiness is worse than a year earlier. How much would this outcome influence your decision about whether or not to have surgery?

Would you still choose **NOT** to have surgery, even if you had this outcome?

☐Yes

☐No

☐Unsure

3.
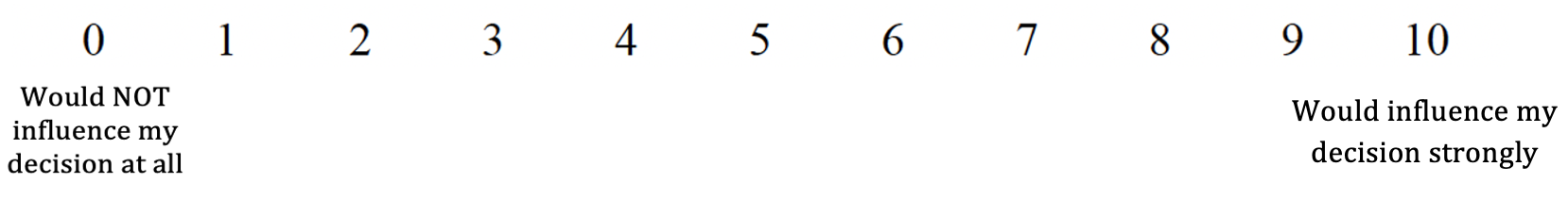
You have a harder time using small objects like buttons or zippers than a year earlier. How much would this outcome influence your decision about whether or not to have surgery?

Would you still choose **not** to have surgery, even if you had this outcome?

☐Yes

☐No

☐Unsure

4.
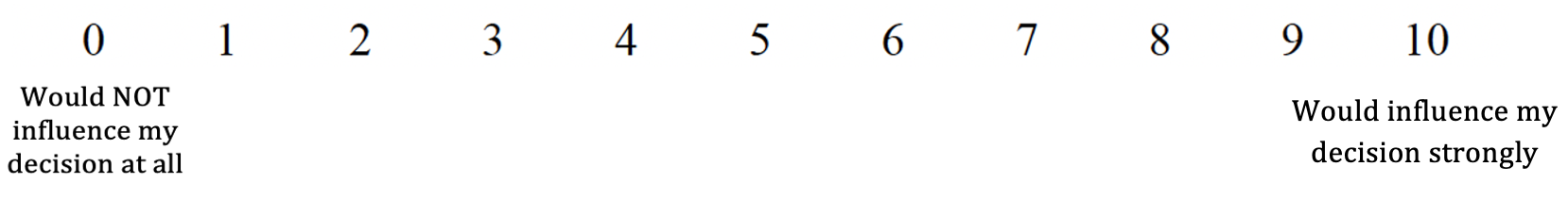
You drop objects more often than you did a year earlier. How much would this outcome influence your decision about whether or not to have surgery?

Would you still choose **not** to have surgery, even if you had this outcome?

☐Yes

☐No

☐Unsure

5. You have more difficulty controlling your bladder than you did a year earlier and have accidents on occasion. How much would this outcome influence your decision about whether or not to have surgery?

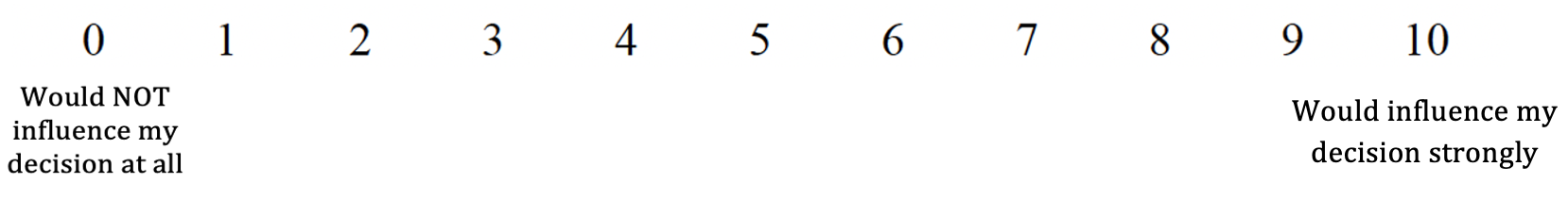

Would you still choose **not** to have surgery, even if you had this outcome?

☐Yes

☐No

☐Unsure

6. You feel more numbness or tingling in your arms than you did a year earlier. How much would this outcome influence your decision about whether or not to have surgery?

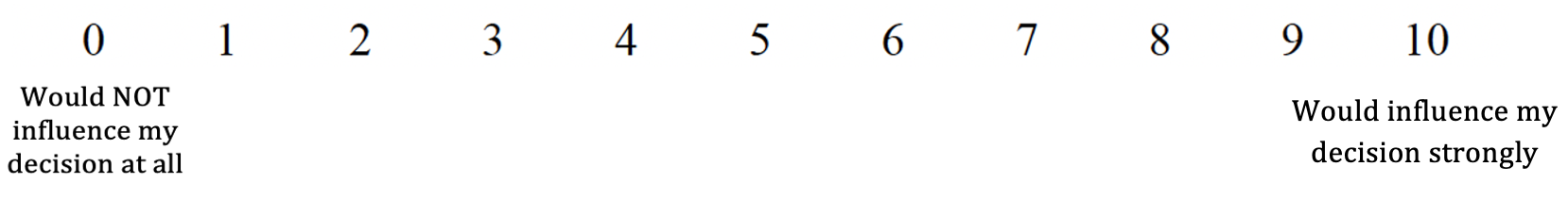

Would you still choose **not** to have surgery, even if you had this outcome?

☐Yes

☐No

☐Unsure

7. You feel more numbness or tingling in your legs than you did a year earlier. How much would this outcome influence your decision about whether or not to have surgery?

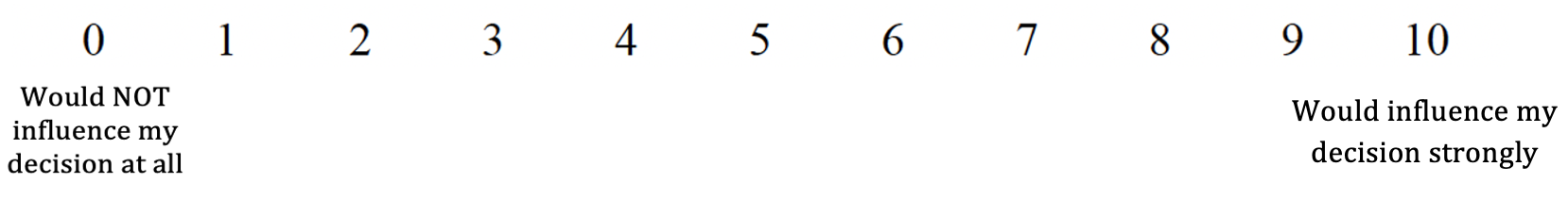

Would you still choose **not** to have surgery, even if you had this outcome?

☐Yes

☐No

☐Unsure

8.
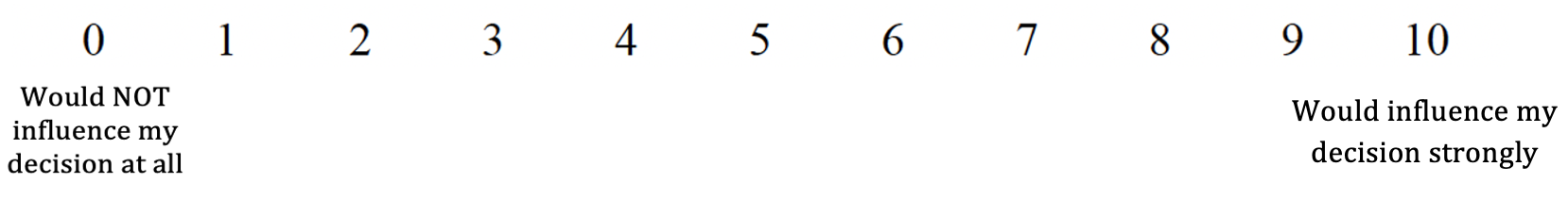
You have increased neck pain that interferes with your sleep than you did a year earlier. How much would this outcome influence your decision about whether or not to have surgery?

Would you still choose **not** to have surgery, even if you had this outcome?

☐Yes

☐No

☐Unsure
